# Generalizability of trial criteria on amyloid-lowering therapy against Alzheimer’s disease to individuals with MCI or early AD in the general population

**DOI:** 10.1101/2024.02.29.24303553

**Authors:** Jacqueline J. Claus, Ilse vom Hofe, Annekee van Ijlzinga Veenstra, Silvan Licher, Harro Seelaar, Frank J. de Jong, Julia Neitzel, Meike W. Vernooij, M. Arfan Ikram, Frank J. Wolters

**Affiliations:** Dept. of Epidemiology, Erasmus MC, Rotterdam, the Netherlands; Dept. of Radiology & Nuclear Medicine, Erasmus MC, Rotterdam, the Netherlands; Dept. of General Practice, Erasmus MC, Rotterdam, the Netherlands; Dept. of Neurology, Erasmus MC, Rotterdam, the Netherlands

**Keywords:** amyloid, clinical trials, monoclonal antibodies, Alzheimer’s disease, generalizability

## Abstract

**Background:** Treatment with monoclonal antibodies against amyloid-β slowed cognitive decline in recent randomized clinical trials in patients with mild cognitive impairment (MCI) and early dementia due to Alzheimer’s disease (AD). However, stringent trial eligibility criteria may affect generalizability of these findings to clinical practice.

**Methods:** We extracted eligibility criteria for trials of aducanumab, lecanemab and donanemab from published reports, and applied these to participants with MCI or early clinical AD dementia from the population-based Rotterdam Study. Participants underwent questionnaires, genotyping, brain MRI, cognitive testing, and cardiovascular assessment. We had continuous linkage with medical records and pharmacy dispensary data. We determined amyloid status using an established and validated prediction model based on age and *APOE* genotype. We assessed progression to dementia within 5 years among participants with MCI, stratified for eligibility.

**Results:** Of 968 participants (mean age: 75 years, 56% women), 779 had MCI and 189 early clinical AD dementia. Across the three drug trials, around 40% of participants would be ineligible because of predicted amyloid negativity. At least one clinical exclusion criterion was present in 76.3% (95% CI; 73.3-79.3) of participants for aducanumab, 75.8% (73.0-78.7) for lecanemab, and 59.8% (56.4–63.3) for donanemab. Criteria that most often led to exclusion were a history of cardiovascular disease (35.2%), use of anticoagulant (31.2%), use of psychotropic or immunological medications (20.4%), history of anxiety or depression (15.9%), or lack of social support (15.6%). One-third of participants were ineligible based on brain MRI findings alone, which was similar across trials and due predominantly to various manifestations of cerebral small-vessel disease. Combining amyloid, clinical, and imaging criteria, eligibility ranged from 9% (7.0-11.1) for aducanumab, 8% (6.2-9.9) lecanemab to 15% (12.4-17.5) for donanemab. Risk of progression to dementia tended to be higher for ineligible than for eligible participants for lecanemab (hazard ratio [95%CI]: 1.64 [0.92-2.91]), aducanumab (HR: 1.17 [0.65-2.12]), and only marginally for donanemab (HR: 1.03 [0.67-1.59]).

**Conclusions:** Findings from recent RCTs reporting protective effects of monoclonal antibodies against amyloid-β are applicable to less than 15% of community-dwelling individuals with MCI or early AD. These findings underline that evidence for drug efficacy and safety is lacking for the vast majority of patients with MCI/AD in routine clinical practice.

## Introduction

Alzheimer’s disease (AD) is the leading cause of dementia, contributing to 38 million cases worldwide.^1^ The search for disease modifying therapy against AD has long been fruitless, with disappointing results from various trials aimed predominantly at the removal of cerebral β-amyloid depositions.^2^ Since 2021, however, three randomized clinical trials of monoclonal antibodies against β-amyloid have shown slowing of cognitive and functional decline in patients with mild cognitive impairment (MCI) and early-stage AD dementia.^3-5^ This led to the conditional approval of aducanumab (June 2021) and regular approval of lecanemab by the United States Food and Drug Administration (FDA, July 2023) and the Japanese New Drug Application (September 2023) as the first disease-modifying therapies.^6-8^ Donanemab has been filed for approval, and a decision by the European Medical Association (EMA) on lecanemab is pending.^9,10^ The approval of these monoclonal antibodies against β-amyloid for clinical use raises the question to which extent findings from trials are generalizable to the wider patient population, in terms of both efficacy and safety.^11^

With regard to efficacy, external validity is of particular relevance for trials against AD, as dementia in the general population is often due to mixed pathology, and roughly half of the explained variance in clinical syndromes of AD dementia is not due to AD-related pathological indices.^12^ Efficacy in patients with pure AD may not apply to patients with comorbid pathology, if the latter did not take part in clinical trials. Indeed, trial eligibility criteria for AD trials preclude certain groups of patients, with criteria ranging from restrictions in age to various types of comorbidities, medication use, and evidence of vascular brain injury on magnetic resonance imaging (MRI). Similarly for safety, restriction of trial enrolment to patients with relatively little comorbidity may underestimate occurrence of adverse effects, like amyloid-related imaging abnormalities (ARIA), in the patient population seen in routine practice.

Three studies have thus far investigated external validity of Alzheimer trials in specialised memory clinic populations. Applying eligibility criteria of aducanumab and lecanemab to these patient populations in Italy and Ireland, 73-99% of participants would have been ineligible for trial inclusion.^13-15^ Similarly, two studies using medical claims data in the United States of America found that over 90% would have been excluded from participation in trials of amyloid-lowering therapy.^16,17^ Population-representative data of European patients are lacking. Moreover, the largest clinical benefit to date has been observed for donanemab in the TRAILBLAZER-ALZ2 trial, but no published studies have yet examined the impact of donanemab trial eligibility on external validity in the general population with MCI and AD dementia.^5^

In this study, we aimed to assess generalizability of trials of efficacious amyloid-lowering drugs by applying trial eligibility criteria to participants with MCI and early dementia due to AD in the population-based Rotterdam Study.

## Methods

### Selection of Trials

On august 14^th^ 2023, we searched the literature for published phase 3 trials of monoclonal antibodies against β-amyloid that met their primary outcome for reducing cognitive and functional decline. Trials included were EMERGE (the high-dose arm of aducanumab), CLARITY-AD (lecanemab), and TRAILBLAZER-ALZ2 (donanemab). Details of included trials are presented in Supplementary Table 1. Trial inclusion and exclusion criteria were obtained from ClinicalTrials.gov and the original trial publications.^3-5^ In case of discrepancy between ClinicalTrials.gov and the published report, we selected criteria from the latter.

### Study Population to assess generalizability of trial findings

The Rotterdam Study is an ongoing population-based cohort study investigating determinants and occurrence of disease in persons aged 40 years and older. The study started in 1990 and now comprises 17,931 individuals living in the Ommoord suburb of Rotterdam, the Netherlands. The design of the Rotterdam Study has been described in detail previously.^18^ In brief, participants are invited for interview and extensive in-person examination at a dedicated research centre every 3-6 years. Routine examinations as part of the Rotterdam Study protocol include standardised questionnaires (e.g., social support, mental health, medication use), cognitive assessment, cardiovascular exams (e.g., blood pressure measurement, electrocardiogram and cardiac ultrasound), psychiatric assessment, venepuncture, and genotyping. From 2005 onward, brain MRI became part of the core study protocol of the Rotterdam Study. For the current study, we included all 968 participants who were diagnosed with MCI (n=779) or early dementia due to clinical manifestation of AD (n=189) during the fourth (2002-2005) and fifth (2009-2013) examination cycle of the study. Participants with dementia included in the current study had to have a dementia diagnosis within 1 year from their study centre visit. From 2005 onwards MRI of the brain was implemented into the core protocol of the Rotterdam Study. In total, 255/454 participants with MCI or early dementia due to AD from the fifth examination round had brain MRI scan available.

The Rotterdam Study has been approved by the Medical Ethics Committee of the Erasmus Medical Centre and by the Ministry of Health, Welfare and Sport of the Netherlands, implementing the Population Studies Act for the Rotterdam Study. All participants provided written informed consent for participation in the study, including permission to obtain information from their treating physicians.

### Ascertainment of mild cognitive impairment and dementia

Participants were screened for dementia at baseline and every 3-6 years during follow-up examinations using the Mini-Mental State Examination (MMSE) and the Geriatric Mental Schedule (GMS) organic level. Those with a MMSE score of <26 or a GMS organic level score of >0 were further examined using the Cambridge Examination for Mental Disorders of the Elderly. Additionally, participants were continuously under surveillance for incident dementia through the electronic linkage between the study database and medical records from general practitioners and the Regional Institute of Outpatients Mental Health Care. The general practitioner functions as a gatekeeper within the Dutch healthcare system, receiving written information of any medical specialists’ consultations of their patients. The final diagnosis of dementia and its most common subtypes was made by a consensus panel led by a neurologist based on the standard criteria for all-cause dementia (DSM-III-R) and clinical Alzheimer’s disease (NINCDS-ADRDA). Follow-up for dementia was completed until January 1, 2020.

Criteria for MCI were based on the Petersen criteria, and included the presence of subjective cognitive complaints and objective cognitive impairment, in the absence of dementia.^19^ A detailed description of our MCI assessment has been published previously.^20^ Subjective cognitive complaints were evaluated by interview, including questions on memory and everyday functioning. We assessed objective cognitive impairment using a cognitive test battery comprising the letter-digit substitution task, Stroop test, verbal fluency test (animal categories), and 15-word verbal learning test based on Rey’s recall of words.^21^ Compound scores were constructed for various cognitive domains, including memory function, information-processing speed, and executive function.^21,22^ We classified persons as objectively cognitively impaired if in any of the cognitive domains, the compound score was at least 1.5 standard deviation lower than expected based on age and education adjusted means, derived from the study population.

### Operationalization of Trial Eligibility Criteria

For each of the trial inclusion and exclusion criteria, we defined an operationalization in the Rotterdam Study. This was done through consensus discussion between two authors (JJC and FJW). A detailed description of the eligibility criteria for each of the three trials, and their operationalization, is provided in Supplementary Tables 2, 3 and 4. For example, history of cardiovascular disease was based on a combination in-person assessment and continuous linkage with medical records for various comorbidities, use of medication was assessed based on pharmacy dispensary data (ATC-codes), and we had in-person questionnaires for social support and screening for anxiety and depression.

### Brain magnetic resonance imaging

Participants underwent scanning on a 1.5-T MRI scanner (GE Healthcare) using a multisequence protocol consisting of T1-weighted, proton density-weighted, fluid-attenuated inversion recovery, and T2-weighted sequences. For brain volumetry, T1-weighted (voxel size 0.49 × 0.49 × 1.6 mm^3^), proton density–weighted (voxel size 0.6 × 0.98 × 1.6 mm^3^), and the fluid-attenuated inversion recovery (FLAIR) (voxel size 0.78 × 1.12 × 2.5 mm^3^) scans were used for automated segmentation of brain tissues, including white matter hyperintensities.^23,24^ In short, a k-nearest neighbor tissue classification algorithm was implemented for quantification of white matter hyperintensities (WMH).^24^ All segmentations were visually inspected, and manually corrected if needed. Severe WMH were defined as a volume of 16.1 milliliter or more, corresponding to a Fazekas score of 3.^25,26^ All scans were appraised by trained research physicians for the presence of cerebral microbleeds (i.e., small round to ovoid hypointense areas on T2*-weighted images), lacunes (i.e., focal cavitating lesions ≥3 and <15 mm), and cortical infarcts.^27^ These ratings were done blinded to clinical data. Furthermore, abnormalities including superficial siderosis, intracranial aneurysms, meningioma, suspected gliomas, cavernous angioma, arachnoid cysts, arteriovenous malformations and dural fistulas were rated by research physicians, and subsequently evaluated by a consultant neuroradiologist.

### Prediction of amyloid status

To assess amyloid positivity, we used a prediction model based on age and *APOE* ε4 allele count.^28^ This model was developed within the Anti-Amyloid Treatment in Asymptomatic Alzheimer’s trial population (A4 Study), and previously showed high discriminative ability to differentiate people with and without brain amyloid as measured by ^18^F-florbetaben PET in the Rotterdam Study population (area under the curve: 0.84 [0.79–0.88]).^28^

### Statistical analyses

Missing data, most notably for social support (17.9%), Parkinson’s disease (due to incomplete assessment of the Unified Parkinson Disease Rating Scale, 7.0%) and anxiety disorder (9.1%), were imputed using 5-fold multiple imputations (“mice” package in R). Details of all imputed variables are described in supplementary table 7.

For each trial, we determined the number of participants who met the eligibility criteria, both overall and per criterion separately. We compared the subset of trial eligible individuals to all individuals with MCI and early AD dementia in the population on several key demographics and participant characteristics, including, age, sex, educational attainment, MMSE, *APOE* ε4 carrier status, MCI or dementia status, cardiovascular disease and antithrombotic medication use. We repeated all analyses in the subsample (n=255) with brain MRI.

Finally, among all individuals with MCI in the primary analyses, we determined 5-year progression to dementia stratified by trial eligibility, by computing hazard ratios (HR) using Cox proportional hazard models.

All analyses were performed in R (version 4.2.1). This study is reported according to the Strengthening the Reporting of Observational Studies in Epidemiology (STROBE) guidelines.^29^

## Results

### Study population

Overall, 968 participants were diagnosed with MCI or early dementia (mean age 75.1 years, 55.2% women), of whom 779 (80.5%) had MCI and 189 (19.5%) clinical AD dementia. Of these, trial-specific entry scores on the MMSE were met by 751 (77.6%) participants for aducanumab (MMSE ≥24), 856 (88.4%) for lecanemab (MMSE ≥22) and 762 (78.7%) for donanemab (MMSE 22-28). Mean (±SD) age of MMSE-eligible participants was similar across trials, varying from 74.0 (±8.0) years for aducanumab, to 74.5 (±8.1) for lecanemab and 75.3 (±8.1) years for donanemab. Little over half of participants were women for all three trials (51.6-55.6%).

### Trial eligibility criteria

Trial eligibility criteria for EMERGE (aducanumab), CLARITY-AD (lecanemab), and TRAILBLAZER-ALZ2 (donanemab) are listed in supplementary tables 2, 3 and 4. Overall, TRAILBLAZER-ALZ2 listed the fewest exclusion criteria. EMERGE was the only trial to exclude persons using platelet inhibiting or anticoagulant medication. CLARITY-AD only included patients with objective impairment in episodic memory, whereas the other two trials did not specify the affected cognitive domain among their inclusion criteria. All trials excluded patients with severe small vessel disease on brain MRI (i.e., severe white matter hyperintensities or >4 microbleeds). EMERGE and CLARITY-AD additionally excluded patients with lacunes or cortical infarct. CLARITY-AD further excluded individuals with intracranial aneurysms, meningioma, cavernous angioma, and arachnoid cysts. A potentially eligible participant has to go through eight different modalities, including cognitive testing, assessment of prior medical history and comedication, BMI measurement, venepuncture, blood pressure measurement, electrocardiogram, brain MRI scan and amyloid-PET scan or lumbar puncture, as well as have an informant/care partner present for the duration of the study.

### Eligibility on the basis of clinical criteria

Around two-thirds of participants met at least one clinical exclusion criterion, somewhat higher for aducanumab (76.3%; 95% CI 73.3-79.3) and lecanemab (75.8%; 73.0-78.7) than for donanemab (59.8%; 56.4–63.3) (Figure 1, Supplementary Table 5). Compared to all individuals with MCI and early AD dementia in the Rotterdam Study, eligible participants were similar with respect to age, sex, educational attainment, *APOE* ε4 carrier status (Table 1). MMSE scores at diagnosis were also similar between groups, although eligible participants were slightly more often at the MCI stage rather than having early dementia. Those eligible for donanemab had more cardiovascular disease (18.3%) compared to those eligible for donanemab (16.4%) and aducanumab (10.1%).

**Figure 1.**
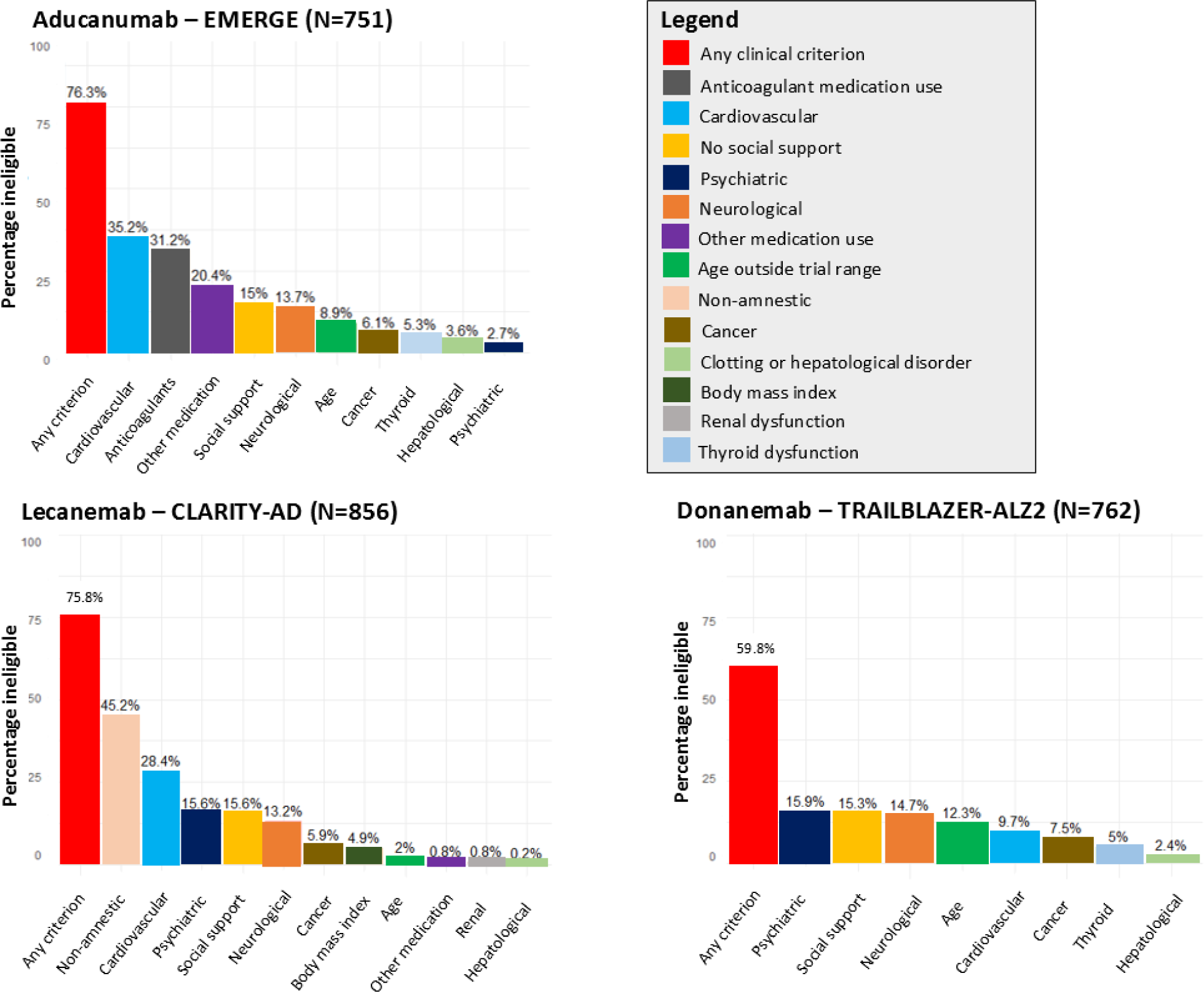
Prevalence of trial eligibility criteria among participants with mild cognitive impairment and early Alzheimer’s Dise ase dementia in the population-based Rotterdam Study. Legend: Trial eligibility criteria among participants with mild cognitive impairment and early Alzheimer’s disease in aducanumab (EMERGE), lecanemab (CLARITY-AD) and donanemab (TRAILBLAZER-ALZ2) trials. Percentages reflect the percentage of participants with individual eligibility criterion, or any criterion.

**Table 1.** Eligibility of all 968 participants with mild cognitive impairment and early Alzheimer’s disease dementia in the Rotte rdam Study.

|  | All participants<br>(n=968) | Aducanumab | Lecanemab | Donanemab |
| --- | --- | --- | --- | --- |
| No. of participants within trial MMSE range |  | 751 | 856 | 762 |
| N (%) eligible based on clinical criteria |  | 178 (23.7) | 207 (24.2) | 306 (40.2) |
| Age, mean (SD) | 75.1 (8.3) | 70.6 (6.7) | 74.3 (8.3) | 73.2 (6.8) |
| Women, n (%) | 534 (55.2) | 90 (50.6) | 124 (59.9) | 161 (52.6) |
| MMSE, mean (SD) | 25.4 (3.3) | 27.1 (1.7) | 26.4 (2.2) | 25.5 (2.2) |
| Low educational attainment<sup>^</sup>, n (%) | 180 (18.6) | 22 (12.4) | 40 (19.3) | 54 (17.8) |
| *APOE* ε4 carriers, n (%) | 301 (32.1) | 63 (35.6) | 55 (27.2) | 113 (37.5) |
| Heterozygote | 270 (28.8) | 56 (31.6) | 50 (24.8) | 100 (33.2) |
| Homozygote | 31 (3.3) | 7 (4.0) | 5 (2.5) | 13 (4.3) |
| Cognitive impairment |  |  |  |  |
| Mild cognitive impairment, n (%) | 779 (80.5) | 160 (89.9) | 178 (86.0) | 258 (84.3) |
| Early dementia, n (%) | 189 (19.5) | 18 (10.1) | 29 (14.0) | 48 (15.7) |
| Cardiovascular disease*, n (%) | 266 (27.5) | 18 (10.1) | 34 (16.4) | 56 (18.3) |
| Antithrombotic medication use, n (%) | 392 (40.5) | 29 (16.3) | 66 (31.2) | 103 (33.7) |
| Antiplatelet | 102 (10.5) | 19 (10.7) | 25 (12.1) | 34 (11.1) |
| Anticoagulant | 290 (30.0) | 0 (0.0) | 41 (19.8) | 69 (22.5) |

Eligibility criteria as applied in the EMERGE trial for aducanumab, CLARITY-AD trial for lecanemab, and TRALBLAZER-ALZ2 trial for donanemab.
No., number; MMSE, Mini-Mental State Examination; SD, standard deviation.
\*indicates composite of previous transient ischemic attack, stroke, myocardial infarction, or coronary artery revascularization procedure.
<sup>^</sup>indicates primary education only
Data were missing for level of education (0.9%) and *APOE* ε4 carriers (3.0%).

Overall, exclusion criteria related to cardiovascular comorbidity had the largest effect on trial eligibility, but its impact differed between trials (Table 1 and Figure 1). For aducanumab, 264/751 (35.2%) participants were excluded because of cardiovascular disease, and 234/751 (31.2%, mostly overlapping) due to use of anticoagulant medication. Cardiovascular disease amounted to 10.7% exclusion of participants for lecanemab and 9.7% for donanemab. Neurological comorbidity and lack of social support were common reasons for ineligibility across trials, affecting 13.2 to 15.6% of potential participants. In trials of lecanemab and donanemab, 16% of people were ineligible based on indication of presence of mental health disorders, due chiefly to positive screening for depression or anxiety disorder. Psychotropic or immunological medication use frequently led to exclusions for aducanumab (20.4%), while for lecanemab only 0.8% met trial-specific exclusions for immunological medication, and donanemab had no such exclusions.

Figure 3 shows the overlap between the different exclusion criteria within participants. Around one third of exclusion criteria across trials occurred in isolation (34% aducanumab, 41% lecanemab, 38% donanemab), as illustrated by the single, unconnected dots in Figure 3. Two thirds of participants who were ineligible due to cardiovascular disease also met one or multiple other exclusion criteria, similar across trials (Figure 3). A notable exception is the presence of non-amnestic MCI (in the absence of memory impairment), which was a common exclusion criterion for lecanemab (391/856, 45.2%), and occurred in isolation in over half of participants meeting this criterion (202/391; 51.7%) (Figure 3).

**Figure 2.**
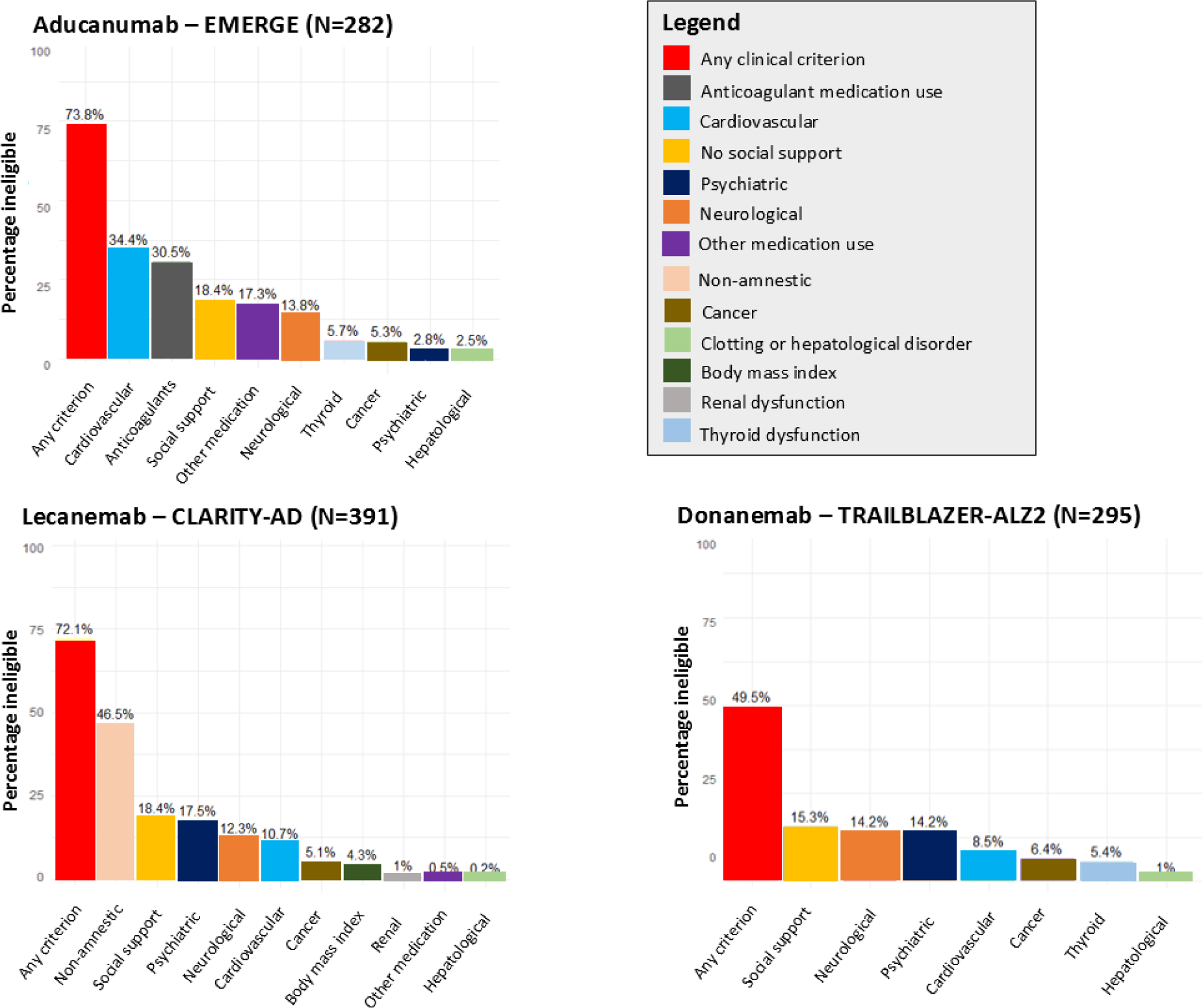
Prevalence of trial eligibility criteria among participants with positive predicted amyloid status and mild cognitive impairment or early Alzheimer’s disease dementia in the population-based Rotterdam Study. Trial eligibility criteria among participants with mild cognitive impairment and early Alzheimer’s disease in aducanumab (EMERGE), lecanemab (CLARITY-AD) and donanemab (TRAILBLAZER-ALZ2) trials. Percentages reflect the percentage of participants with individual eligibility criterion, or any criterion.

**Figure 3.**
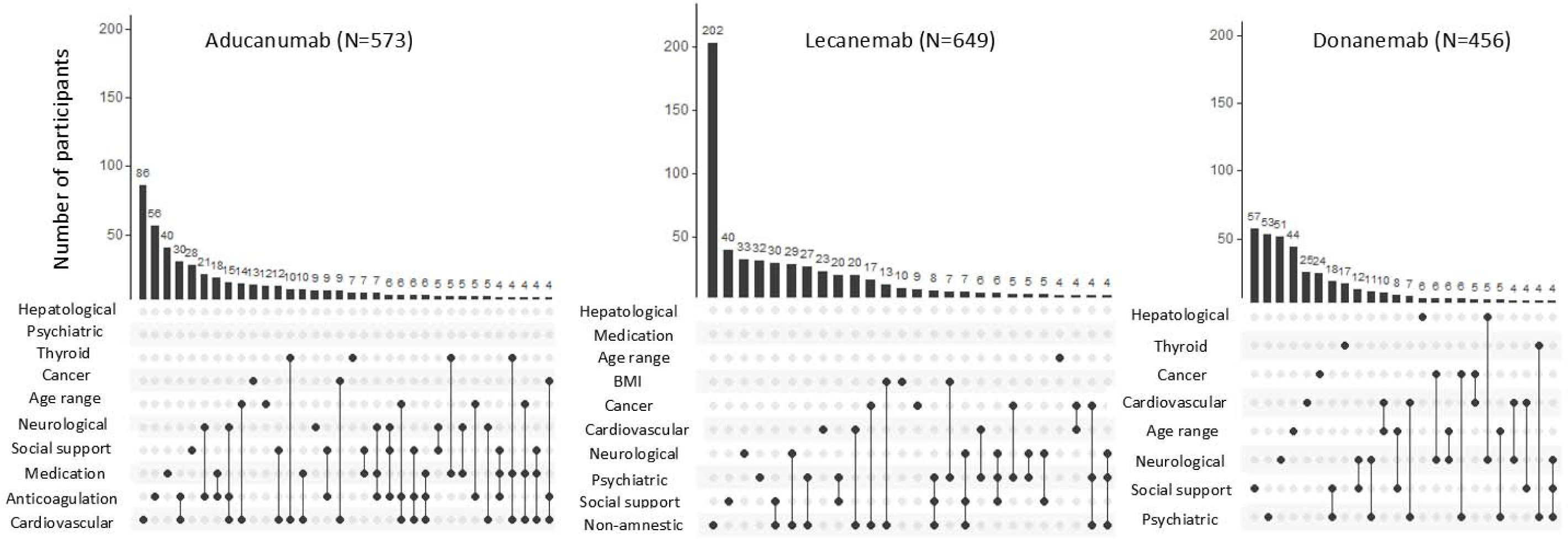
Concurrence of eligibility criteria for aducanumab, lecanemab and donanemab. Legend: These intersection diagrams show the patterns of co-occurrence of different eligibility criteria. Rows represent the types of eligibility criteria and columns represent their combinations. All criteria that are part of a given combination are shown as black dots connected by a vertical black line. A single dot without a line implies the symptom occurred in isolation. The number of participants with a given combination of criteria is shown as a vertical bar on top of the matrix. The graph is truncated at a minimum of four participants per combination. BMI indicates body mass index.

### Impact of (predicted) amyloid positivity

Amyloid status was predicted negative in around 40% of all participants with MCI and early clinical AD dementia, similar across trials (aducanumab 42.3%, lecanemab 40.4%, donanemab 37.9%). When adding amyloid negativity to the aforementioned clinical eligibility criteria, the percentage of persons who were ineligible increased from 76.3 to 88.4% for aducanumab, from 75.8 to 86.0% for lecanemab, and from 62.3 to 76.4% for donanemab. Conversely, among all age and MMSE eligible Rotterdam Study participants with positive predicted amyloid status, application of clinical exclusion criteria left 73.8% of participants ineligible for aducanumab, 78.2% for lecanemab, and 49.5% ineligible for donanemab (Figure 2 and Supplementary Table 6).

### MRI criteria

Of all 968 included participants, 255 had an available brain MRI scan close to MCI/early AD diagnosis (mean interval: 2.3 years). The most common imaging abnormalities were of presumed vascular nature, with severe white matter hyperintensities in 24.6%, cortical infarcts in 10.6%, ≥2 lacunes in 7.5%, and >4 microbleeds in 3.1% of participants. Around one third of participants would be ineligible for trial participation on the basis of imaging criteria alone, which was similar across trials (Figure 4).

**Figure 4.**
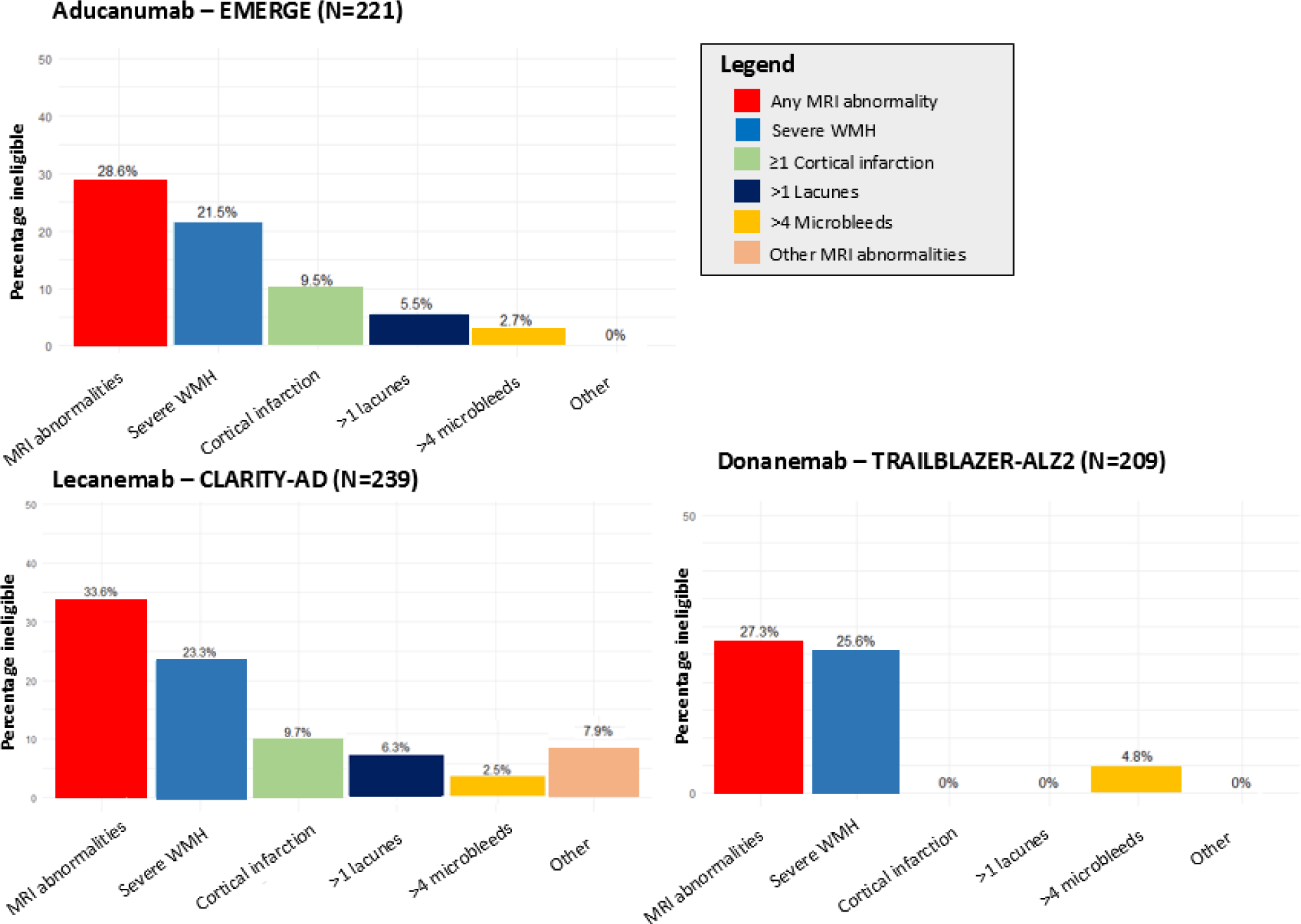
Impact of MRI criteria on trial eligibility. Legend: Prevalence of trial eligibility criteria for brain magnetic resonance imaging (MRI) scan among participants with mild cognitive impairment and Alzheimer’s disease dementia in aducanumab (EMERGE), lecanemab (CLARITY-AD) and donanemab (TRAILBLAZER-ALZ2) trails. Percentages reflect the percentage of participants with individual eligibility criterion, and MRI abnormalities reflect the presence of any of the MRI criteria. MRI indicates magnetic resonance imaging, WMH indicates white matter hyperintensities.

Eligibility based on clinical criteria was similar in the MRI subsample compared to the overall sample. Compared to the entire sample, participants who were eligible based on clinical and MRI criteria were slightly younger, less educated and more often carrier of the *APOE ε*4 allele (Table 2). A combination of all clinical criteria, MRI criteria, and predicted amyloid negativity resulted in trial ineligibility of 91.1% (95% CI, 88.9-93.0) for aducanumab, 91.6% (90.1-93.8) for lecanemab and 85.2% (82.5-87.6) for donanemab.

**Table 2.** Baseline characteristics for the overall Rotterdam Study cohort with mild cognitive impairment and early Alzheimer’s disease dementia and according to their eligibility for published trials.

|  | All participants<br>(n=255) | Aducanumab<sup>1</sup> | Lecanemab<sup>2</sup> | Donanemab<sup>3</sup> |
| --- | --- | --- | --- | --- |
| No. of participants with available MRI scan |  | N = 221 | N = 239 | N = 209 |
| N (%) eligible based on MRI criteria |  | 158 (71.5) | 159 (66.5) | 152 (72.7) |
| N (%) eligible based on clinical + MRI criteria |  | 49 (22.2) | 39 (16.3) | 64 (30.6) |
| Age, mean (SD) | 74.0 (7.3) | 70.0 (6.4) | 69.6 (6.7) | 71.5 (5.9) |
| Women, n (%) | 113 (44.3) | 24 (49.0) | 18 (46.2) | 29 (45.3) |
| MMSE, mean (SD) | 26.3 (2.4) | 26.9 (1.6) | 26.9 (1.5) | 26.2 (1.8) |
| Low educational attainment, n (%) | 33 (13.0) | 6 (12.2) | 7 (17.9) | 10 (15.6) |
| APOE ε4 carriers, n (%) | 76 (30.6) | 19 (38.7) | 15 (39.5) | 26 (41.3) |
| Heterozygote | 68 (27.4) | 17 (34.7) | 14 (36.8) | 25 (39.7) |
| Homozygote | 8 (3.2) | 2 (4.1) | 1 (2.6) | 1 (1.6) |
| Cognitive impairment |  |  |  |  |
| Mild cognitive impairment, n (%) | 228 (89.4) | 45 (91.8) | 37 (94.9) | 57 (89.1) |
| Early clinical AD dementia, n (%) | 27 (10.6) | 4 (8.2) | 2 (5.1) | 7 (10.9) |
| Cardiovascular disease<sup>^</sup>, n (%) | 74 (29.0) | 7 (14.3) | 4 (10.3) | 8 (12.5) |
| Antithrombotic medication use, n (%) | 113 (44.3) | 9 (18.4) | 8 (20.5) | 10 (16.5) |
| Antiplatelet | 34 (13.3) | 9 (18.4) | 7 (17.9) | 9 (14.1) |
| Anticoagulant | 79 (31.0) | 0 (0.0) | 1 (2.6) | 1 (1.6) |

<sup>1</sup> Eligibility criteria as applied in EMERGE trial. <sup>2</sup> Eligibility criteria as applied in CLARITY-AD trial. <sup>3</sup> Eligibility as applied in TRALBLAZER-ALZ2 trial. No., number; MMSE, Mini-Mental State Examination; SD, standard deviation. <sup>^</sup> indicates composite of previous transient ischemic attack, stroke, myocardial infarction, percutaneous coronary intervention or coronary artery bypass grafting. Data were missing for level of education (0.8%) and APOE ε4 carriers (2.7%).

### Dementia progression

During 5-year follow-up, progression to dementia was observed in 91 Rotterdam Study participants with MCI. Participants with MCI who were ineligible based on clinical criteria tended to have higher risk of progressing to dementia than eligible participants for lecanemab (HR: 1.64, 95% CI: 0.92 – 2.91), aducanumab (HR: 1.17, 95% CI: 0.65 – 2.12) and only marginally for donanemab (HR: 1.03, 95% CI: 0.67 – 1.59).

## Discussion

In this population-based study, findings from recent RCTs of monoclonal antibodies against amyloid-β are applicable to less than 15% of persons with MCI and early clinical AD dementia. This percentage was somewhat higher for donanemab (15%) than for aducanumab and lecanemab (8-9%). Trial ineligibility was commonly due to comorbid vascular pathology, either clinically or on brain MRI, neurological or psychiatric comorbidity, lack of social support, and concurrent anticoagulant or psychotropic medication use.

Thus far, no published studies have assessed eligibility criteria for the TRAILBLAZER-ALZ2 trial of donanemab, which had somewhat less stringent criteria compared to aducanumab and lecanemab. Eligibility for donanemab is slightly higher compared to aducanumab and lecanemab, due to more liberal inclusion of patients with cardiovascular comorbidity, but still limited at 15%. The observed eligibility of around 10% for aducanumab and lecanemab in this population-representative European sample is in accordance with two previous population-based studies from the United States.^16,17^ Clinic-based studies from Italy and Ireland reported eligibility between 1% and 27%.^13-15^

Ineligibility in our study, as in previous reports, was mainly due to comorbid cardiovascular and cerebrovascular disease.^16,17^ The limited inclusion of patients with vascular comorbidity underlines the need for caution and careful monitoring of drug safety if prescribing anti-amyloid-β antibodies to a yet untested patient population. Moreover, in patients with a combination of AD and vascular disease, cognitive impairment is often in part a consequence of cerebrovascular pathology,^30^ and observed efficacy of monoclonal antibodies against amyloid-β in patients with relatively pure AD may not apply to those individuals.

Stringent eligibility criteria in AD trials serve in part to reduce safety and tolerability concerns.^15^ Patients excluded from clinical trials, particularly those with cardiovascular comorbidity, may face a conceivably higher risk of side effects and adverse events. Although individuals using antithrombotic medications did not demonstrate an elevated risk of ARIA in the trial of lecenamab,^31^ these findings might not apply to non-participants with higher frailty or cardiovascular comorbidity. Moreover, any benefits of monoclonal antibodies against cognitive decline in patients with AD who are at high risk of ischemic stroke, should be carefully weighed against the contraindication for intravenous tissue plasminogen activator in the event of a stroke whilst on monoclonal antibodies against amyloid-β.^32^ Risk of ARIA is especially high in homozygote *APOE* ε4 carriers, which prompts special consideration for this group in the FDA’s appropriate use criteria of lecanemab.^33,34^ In our overall study sample, *APOE* ε4 carriership was somewhat lower than in clinical trials and a prior eligibility study from the U.S.,^3-5,17^ but among those with predicted amyloid positivity, *APOE* ε4 carriership was consistent at around 80%.

At present, a large number of patients with cognitive impairment defers from seeking medical attention until a fairly advanced stage of the disease. With the clinical availability of medication against AD, it is expected that a substantial number of people with MCI will attend their physician sooner, in the hope of disease-modifying treatment. Adequate preparation is imperative, and includes the necessary diagnostics for determining amyloid status, monitoring safety, and estimating the contribution of amyloid – relative to other contributing causes – to cognitive decline within a single patient.^35,36^ The current study, along with the representative sample of U.S. citizens,^17^ offers guidance as to the group of patients for which there is some evidence of treatment efficacy, and those for which there is none as of yet. Even if treatment labels would strictly follow trial eligibility criteria, the absolute number of patients eligible for treatment will pose a challenge to healthcare systems.

The current study is strengthened by its large, population representative sample with detailed information of demographics, clinical characteristics, and brain imaging. When interpreting our findings, however, some limitations need to be acknowledged. First, brain MRI was available only in a subsample of the Rotterdam Study cohort. The applied prediction algorithm for amyloid positivity may have led to some misclassification of amyloid status. However, the prediction algorithm was derived externally and validation in the Rotterdam Study population (using amyloid-PET) achieved high discriminatory value.^28^ Amyloid-PET is available in a subsample of the Rotterdam Study, but included only seven and thus too few participants with MCI and early dementia to compute meaningful eligibility numbers. Second, operationalization of the eligibility criteria in the Rotterdam Study was at times intrinsically hard to define (e.g., “the participants need to be in good health as determined by the investigator”), or – incidentally– unavailable in the Rotterdam Study (i.e., childbearing potential and sensitivity to PET tracers). Furthermore, availability of an informant/caregiver in the Rotterdam Study might differ from that in clinical practice. Third, we did not include persons who developed dementia within the study period but did not attend the research centre within 1 year from diagnosis. This might have led to some underestimation of ineligibility, as these persons were older and most likely had more comorbid pathology or were “not in good health as determined by the investigator”. Last, the Rotterdam Study consists of a predominantly White population, and generalizability to non-White populations remains to be determined.^37^

In conclusion, findings from recent RCTs reporting protective effects of monoclonal antibodies against amyloid-β are applicable to less than 15% of community-dwelling individuals with MCI or early AD. These findings underline that evidence for drug efficacy and safety is still lacking for the majority of patients with AD in the community.

## Data Availability

Data can be obtained upon request. Requests should be directed towards the management team of the Rotterdam Study, which has a protocol for approving data requests. Because of restrictions based on privacy regulations and informed consent of the participants, data cannot be made freely available in a public repository. FJW had full access to the data in the study and takes responsibility for data integrity and accuracy of data analysis.

