## Supplementary material for "Generalizability of trial criteria on amyloid-lowering therapy against Alzheimer’s disease to individuals with MCI or early AD in the general population"

**Supplementary Table 1: Study Characteristics of Included Trials**

| Clinical Trial Number | Trial | Drug | Study Phase | Study Start | Study end | Number of Participants |
| --- | --- | --- | --- | --- | --- | --- |
| NCT 02484547 | EMERGE | Aducanumab | 3 | Sep-15 | Aug-19 | 1643 |
| NCT 03887455 | CLARITY-AD | Lecanemab | 3 | Mar-19 | Sep-27 | 1906 |
| NCT04437511 | TRAILBLAZER-ALZ2 | Donanemab | 3 | Jun-2020 | Apr-2023 | 1800 |

| **Supplement Table 2. EMERGE (aducanumab) Clinical Trial Inclusion and Exclusion Criteria and Operationalization in the Rotterdam Study** | |
| --- | --- |
| **Clinical Trial Inclusion Criteria** | **Operational Study Definition** |
| Age 50 to 85 Years. | Age between 50 and 85 years at date of research center visit, based on date of birth from municipality data.  Note: as by the definition of mild cognitive impairment in the Rotterdam Study, no participants with mild cognitive impairment were aged younger than 60 years during the research center visit. |
| Must meet all of the following clinical criteria for MCI due to AD or mild AD according to NIA-AA criteria, and must have:   - A CDR global score of 0.5. - An RBANS score of 85 or lower indicative of objective cognitive impairment (based upon the Delayed Memory Index score). - An MMSE score between 24 and 30 (inclusive). | Diagnosis of mild cognitive impairment (MCI) at study center visit, as defined as presence of i) subjective cognitive complaints, ii) objective cognitive impairment, and iii) absence of dementia. Details of MCI diagnosis in the Rotterdam Scan Study is described elsewhere^1^.    Diagnosis of Alzheimer’s disease within one year prior to or one year after research center visit according to NINCDS-ADRDA criteria, continuously monitored through linkage with computerized medical records and discussed through a panel with an expert neurologist^2^.  Mini-mental state exam score (MMSE) of ≥ 24 assessed during home interview. |
| Must have a positive amyloid PET scan. Previously obtained PET scan (within 12 months of Screening) is permissible for subjects not participating in the amyloid PET sub-study. Previous PET scan images must be submitted to the central imaging vendor to confirm study inclusion criteria are met. | Predicted positive amyloid positive emission tomography (PET) status assessed based on a in the Rotterdam Study validated prediction model (area under the curve = 0.84), including age and apolipoprotein (APOE) ε4 allele count (0,1,2)^3^. |
| Consent to APOE genotyping. | All participants provided informed consent and authorization to partake in the Rotterdam Study and to monitor medical records, all participants were in full ability to understand the purpose and risks of the study. |
| Has one informant/care partner who, in the investigator’s opinion, has frequent and sufficient contact with the subject as to be able to provide accurate information about the subject’s cognitive and functional abilities. The informant/care partner must minimally be available by phone to provide information to the investigator and study staff about the subject and agrees to attend in person clinic visits that require partner input for scale completion. An informant/care partner should be available for the duration of the study, and the use of the same informant/care partner for the duration of the study is encouraged. | Has sufficient social support, as assessed based on a questionnaire “Do you have someone who can give you help when needed?”, answered with “yes”. Answers “no” and “maybe” were defined as insufficient social support. |
| Ability to understand the purpose and risks of the study and provide signed and dated informed consent and authorization to use confidential health information in accordance with national and local subject privacy regulations. | All participants provided informed consent and authorization to partake in the Rotterdam Study and to monitor medical records, all participants were in full ability to understand the purpose and risks of the study. |
| All women of childbearing potential and all men must practice highly effective contraception during the study and for 24 weeks after their last dose of study treatment. | All women included were aged 50 years or older and are presumed not to be at childbearing potential. Criteria for men were not available in the Rotterdam Study. |
| Must have at least 6 years of education or work experience to exclude mental deficits other than MCI or mild AD. | All participants in the Rotterdam Study had at least 6 years of education according to the International Standard Classification of Education (ISCED) of the United Nations of Educational, Scientific and Cultural Organization (UNESCO)^4^. |
| Apart from a clinical diagnosis of early AD, the subject must be in good health as determined by the investigator, based on medical history and screening assessments. | Full description of operationalization of specific criteria can be found in exclusion criteria. |

| **Clinical Trial Exclusion Criteria** | **Operational Study Definition** |
| --- | --- |
| Any uncontrolled medical or neurological/neurodegenerative condition (other than AD) that, in the opinion of the investigator, might be a contributing cause of the subject’s cognitive impairment (e.g., substance abuse, vitamin B_12_ deficiency, abnormal thyroid function, stroke or other cerebrovascular condition, Lewy body dementia, frontotemporal dementia, head trauma). | Prevalent Parkinson’s disease and stroke at research center visit, assessed by interview and verified using medical records.  History of Transient Ischemic Attack (TIA) within 12 months prior to research center visit, as assessed by interview and verified using medical records.  Prevalent brain cancer at research center visit, assessed by medical records and through linkage with a nationwide registry of histo- and cytopathology in the Netherlands, Pathologisch-Anatomisch Landelijk Geautomatiseerd Archief (PALGA).  Prevalent multiple sclerosis, as assessed by medical records.  Presence of epilepsy at research center visit, based on a questionnaire “Did you experience an episode of epilepsy since the last research visit?”.  Presence of abnormal thyroid function at home visit, as assessed based on a questionnaire “Since the last research visit, did you experience any disease concerning the thyroid that needed treatment from the general practitioner or a specialist?”. |
| Transient ischemic attack or stroke or any unexplained loss of consciousness within 1 year prior to Screening. | History of stroke or TIA within 12 months prior to research center visit, as assessed by interview and by continuous linkage with medical records. |
| Clinically significant unstable psychiatric illness (e.g., uncontrolled major depression, uncontrolled schizophrenia, uncontrolled bipolar affective disorder) within 6 months prior to Screening. | History of major depression or bipolar disorder within 6 months prior to research center visit, assessed using medical records. |
| History of unstable angina, myocardial infarction, chronic heart failure (New York Heart Association Class III or IV), or clinically significant conduction abnormalities (e.g., unstable atrial fibrillation) within 1 year prior to Screening. | History of heart failure, coronary heart disease (including myocardial infarction, percutaneous coronary angioplasty and coronary artery bypass grafting) or incident atrial fibrillation within 12 months prior to research center visit, monitored using medical records. |
| Brain MRI performed at Screening (per centrally read MRI) that shows evidence of any of the following:   - Acute or sub-acute hemorrhage. - Prior macrohemorrhage (defined as >1 cm in diameter on T2* sequence) or prior subarachnoid hemorrhage unless it can be documented that the finding is not due to an underlying structural or vascular abnormality (i.e., finding does not suggest subject is at risk of recurrent hemorrhage). - Greater than 4 microhemorrhages (defined as £ 1 cm in diameter on T2* sequence). - Cortical infarct (defined as >1.5 cm in diameter; irrespective of anatomic location). - >1 lacunar infarct (defined as <1.5 cm in diameter). - Superficial siderosis. - History of diffuse white matter disease as defined by a score of 3 on the age-related white matter changes scale. - Any finding that, in the opinion of the investigator, might be a contributing cause of subject’s dementia, might pose a risk to the subject, or might prevent a satisfactory. - MRI assessment for safety monitoring. | Brain magnetic resonance imaging (MRI) shows no evidence of > 1 lacunar infarcts, cerebellar infarctions, cortical infarctions, > 4 microbleeds or superficial siderosis based on visual ratings. A detailed protocol of the Rotterdam Scan Study is described elsewhere^5^.  Fazekas score of 3 based on volumetric segmentations (>16.1ml of white matter hyperintensities)^5^. |
| Recent history (within 1 year of Screening) of alcohol or substance abuse as determined by the investigator, a positive urine drug (due to nonprescription drug) or alcohol test at Screening, or use of cannabinoids (prescription or recreational). | For the fourth examination round research visits, the presence of alcohol abuse based on the CAGE questionnaire^6^.  For the fifth visits, assessed by a questionnaire “How many units of alcohol do you consume during one week?” answered with ≥ 6. |
| History of bleeding disorder or predisposing conditions, blood clotting or clinically significant abnormal results on coagulation profile at Screening, as determined by the investigator. | History of deep vein thrombosis or pulmonary embolism, assessed by in-person questionnaires during home interview: “Since the last research visit, did you experience deep vein thrombosis of the leg?” and “Since the last research visit, did you experience pulmonary embolism?”. |
| Presence of diabetes mellitus that, in the judgment of the investigator, cannot be controlled or adequately managed. | Prevalent or incident diabetes at time of research center visit determined by the use of blood glucose-lowering therapy and uncontrolled by fasting serum glucose levels of ≥ 8.0 millimole/L at research center visit despite use of blood-glucose lowering therapy. |
| Clinically significant 12-lead ECG abnormalities, as determined by the investigator. | Presence of second or third degree atrioventricular (AV) block based on MEANS interpretation, assessed by research physicians using electrocardiogram (ECG) from the research center visit^7^. |
| Uncontrolled hypertension defined as: average of 3 systolic blood pressure [SBP]/diastolic blood pressure [DBP] readings > 165 mmHg and/or > 100 mmHg at Screening (blood pressure measurements exceeding these limits may be repeated as warranted by the investigator, but values must be within the specified limits for the subject to be eligible for the study), or persistent SBP/DBP readings > 180 mmHg and/or > 100 mmHg 3 months prior to randomization (Day 1) that, in the opinion of the investigator, are indicative of chronic uncontrolled hypertension. | Systolic blood pressure > 165 mmHg or diastolic blood pressure > 100 mmHg, assessed by the mean of two blood pressure assessment at the right arm during the research center visit. |
| History of malignancy or carcinoma. The following exceptions may be made after discussion with the Sponsor:   - Subjects with cancers in remission more than 5 years prior to Screening. - Subjects with a history of excised or treated basal cell or squamous carcinoma of the skin. - Subjects with localized prostate cancer with treatment cycles that completed at least 6 months prior to Screening. | History of cancer (solid and hematological, with exclusion of non-melanoma skin cancer) within 5 years prior to research center visit, as assessed by medical records and through linkage with a nationwide registry of histo- and cytopathology in the Netherlands, PALGA. |
| History of seizure within 10 years prior to Screening. | Screening of medical records in combination with structured interview assessing a history of epilepsy within 5-10 years prior to research center visit (i.e., “Did you experience an episode of epilepsy  since the last research visit?”) |
| Indication of impaired liver function as shown by an abnormal liver function profile at Screening (e.g., repeated values of aspartate aminotransferase [AST] and alanine aminotransferase [ALT] ≥ 2 × the upper limit of normal). | During the fifth research center visit, positive test result screening for the presence of indication of impaired liver function, assessed by aspartate aminotransferase and alanine aminotransferase levels > 90. Not available for the fourth research center visit. |
| History or evidence of an autoimmune disorder considered clinically significant by the investigator or requiring chronic use of systemic corticosteroids or other immunosuppressants. | Use of immunosuppressant medication (ATC-code: L04A) and corticosteroids (ATC-code: H02AB), based on pharmacy dispensing records. |
| Clinically significant systemic illness or serious infection (e.g., pneumonia, septicemia) within 30 days prior to or during Screening. | Not available in the Rotterdam Study. |
| History of or known seropositivity for human immunodeficiency virus (HIV). | Not available in the Rotterdam Study. |
| History of or positive test result at Screening for hepatitis C virus antibody or hepatitis B virus (defined as positive for both hepatitis B surface antigen AND hepatitis B core antibody). | During the fifth research center visit, positive test result screening for hepatitis C virus antibody or hepatitis B virus, based on level of ≥ 1.0 in the Elecsys Anti-HCV II and the HbsAg immunoassays ^8^. Not available for the fourth research center visit. |
| History of severe allergic or anaphylactic reactions, or history of hypersensitivity to any of the inactive ingredients in the drug product (refer to the IB for information on the clinical formulation). | Not available in the Rotterdam Study. |
| Any other medical conditions (e.g., renal disease) that are not stable or controlled, or, which in the opinion of the investigator, could affect the subject’s safety or interfere with the study assessments. | Not available in the Rotterdam Study. |
| Any medications that, in the opinion of the investigator, may contribute to cognitive impairment, put the subject at higher risk for AEs, or impair the subject’s ability to perform cognitive testing or complete study procedures. | Current use of benzodiazepines (ATC-code: N05BA, N05CD), antipsychotics (ATC-code N05AA, N05AB, n05AC, N05AD, N05AF, N05AG, N05AH, 05AL, N05AN, N05AX), tricyclic antidepressants (ATC-code: N06AA), tolterodine (ATC-code G04BD07) and anti-epileptic medication (ATC-code: N03A) at research center visit, based on pharmacy dispensing records. |
| Use of allowed chronic medications at doses that have not been stable for at least 4 weeks prior to Screening Visit 1 and during Screening up to Study Day 1, or use of AD medications (including but not limited to donepezil, rivastigmine, galantamine, tacrine, and memantine) at doses that have not been stable for at least 8 weeks prior to Screening Visit 1 and during Screening up to Study Day 1. | Not available in the Rotterdam Study. |
| Use of medications with platelet anti-aggregate or anti-coagulant properties (the use of aspirin at a prophylactic dose [≤ 325 mg daily] is allowed). | Current use of blood thinners (ATC-code: B01A), including fenprocoumon, acenocoumarol, dalteparin, nadroparin, tinzaparin, clopidogrel, dipyridamole, carabasalatecalcium, prasugrel, ticagrelor, clopidogre/acetylsalicylacid, dabigratran, rivaroxaban and apixaban at research center visit, based on pharmacy dispensing records. |
| Use of illicit narcotic medication. | Not available in the Rotterdam Study. |
| Vaccinations within 10 days prior to randomization (Day 1). | Not available in the Rotterdam Study. |
| Participation in any active immunotherapy study targeting Ab unless documentation of receipt of placebo is available. | Not available in the Rotterdam Study. |
| Participation in any passive immunotherapy study targeting Ab within 12 months of Screening unless documentation of receipt of placebo is available. | Not available in the Rotterdam Study. |
| Participation in any study with purported disease-modifying effect in AD within 12 months prior to Screening unless documentation of receipt of placebo is available. Subjects who developed `-E during a previous disease-modifying trial should be excluded. | Not available in the Rotterdam Study. |
| Participation in a previous study with aducanumab (subject is eligible if he/she did not receive active aducanumab). | Not available in the Rotterdam Study. |
| Contraindications to having a brain MRI (e.g., pacemaker; MRI-incompatible aneurysm clips, artificial heart valves, or other metal foreign body; claustrophobia that cannot be medically managed). | Presence of a contraindication for MRI. |
| Contraindication to having a PET scan (e.g., inability to lie flat or still for the duration of the scan) or intolerance to previous PET scans (i.e., previous hypersensitivity reactions to any PET ligand or imaging agent, failure to participate in and comply with previous PET scans). | Not available in the Rotterdam Study. |
| A negative PET scan result with any amyloid-targeting ligand within 6 months prior to Screening. | Not available in the Rotterdam Study. |
| Have had or plan exposure to experimental radiation within 12 months prior to Screening such that radiodosimetry limits would be exceeded by participating in this study. | Not available in the Rotterdam Study. |
| For subjects who consent to LP, any contraindications to having a LP (e.g., platelet count < 100,000/μL, lumbar spine deformity). Any symptoms caused by or related to the optional LP during Screening must be resolved prior to randomization (Day 1). Subjects may still participate in the overall study even if participation in the optional LP portion is contraindicated. | Not determined since subjects may still participate in the overall study. |
| Female subjects who are pregnant or currently breastfeeding. | All women included were aged 50 years or older and are presumed not to be at childbearing potential. Criteria for men were not available in the Rotterdam Study. |
| Previous participation in this study. Subjects who fail Screening will be permitted to be rescreened once at the Sponsor’s discretion, except those who fail due to PET, MMSE, CDR global score > 0.5, hepatitis B or C, or abnormal MRI findings. (Subjects who fail Screening due to a CDR global score of 0 may be rescreened; such subjects will be allowed to repeat the screening CDR assessment after 6 months.). | Not available in the Rotterdam Study. |
| Subject currently living in an organized care facility with extensive intervention and/or support of daily living activities. | Not available in the Rotterdam Study. |
| Blood donation (≥ 1 unit) within 1 month prior to Screening. | Not available in the Rotterdam Study. |
| Inability to comply with study requirements. | Not available in the Rotterdam Study. |
| Other unspecified reasons that, in the opinion of the investigator or Biogen, make the subject unsuitable for enrollment. | Not available in the Rotterdam Study. |

| **Supplement table 3. CLARITY-AD (lecanemab) Clinical Trial Inclusion and Exclusion Criteria and Operationalization in the Rotterdam Study** | |
| --- | --- |
| **Clinical Trial Inclusion Criteria** | **Operational Study Definition** |
| Age ≥ 50 Years and ≤ 90 Years. | Age between 50 and 90 years at date of research center visit, based on date of birth from municipality data.  Note: as by the definition of mild cognitive impairment in the Rotterdam Study, no participants with mild cognitive impairment were aged younger than 60 years during the research center visit. |
| Diagnosis MCI due to Alzheimer’s disease (NIA-AA core clinical criteria, CDR 0.5 or greater, report history of subjective memory decline with gradual onset and slow progression over 1 year) or Mild Alzheimer’s disease dementia (NIA-AA core clinical criteria probable AD, CDR 0.5 to 1.0 and memory Box score 0.5 or greater). | Diagnosis of MCI at study center visit, as defined as presence of i) subjective cognitive complaints, ii) objective cognitive impairment, and iii) absence of dementia. Details of MCI diagnosis in the Rotterdam Scan Study is described elsewhere^1^.    Diagnosis of Alzheimer’s disease within one year prior to or one year after research center visit according to NINCDS-ADRDA criteria, continuously monitored through linkage with computerized medical records and discussed through a panel with an expert neurologist^2^. |
| MMSE score greater than or equal to 22 at Screening and Baseline and less than or equal to 30 at Screening and Baseline. | MMSE score of ≥ 22 assessed during home interview. |
| Objective impairment in episodic memory as indicated by at least 1 standard deviation below age-adjusted mean in Wechsler Memory Scale IV-Logical Memory subscale II.  a. ≤ 15 for age 50 to 64 years  b. ≤ 12 for age 65 to 69 years  c. ≤ 11 for age 70 to 74 years  d. ≤ 9 for age 75 to 79 years  e. ≤ 7 for age 80 to 90 years | Objective impairment in verbal memory as indicated by a score at least 1 standard deviation below age- and education adjusted means based on the population from the fourth examination round of the Rotterdam Study. This round was taken as the norm population, since this was a generally representative population with the largest dataset. |
| Positive biomarker for brain amyloid pathology   a. PET assessment of imaging agent uptake into brain. Note: amyloid PET screens will be performed according to local regulatory guidelines and thus may be restricted for those subjects who are not suitable for lumbar puncture (LP) to obtain CSF for testing of eligibility. b. CSF assessment of t-tau/Aβ[1-42].   NOTE: Subjects may consent to both the PET and CSF assessments, but to confirm eligibility, a positive amyloid result is needed in only 1 of the 2 procedures (ie, the subject will be eligible even if 1 of the 2 results does not meet its eligibility criterion). Subjects who consent to amyloid PET or CSF at Screening for the purposes of eligibility are not required to participate in the amyloid PET, tau PET, or CSF longitudinal substudies. Use of a historical amyloid positive PET (conducted within 12 months before the planned date of randomization) is acceptable for determination of eligibility but will not suffice for the baseline assessment if the subject wishes to consent to the amyloid PET longitudinal Clinical Study Protocol BAN2401-G000-301 substudy. The historical imaging data must be made available to the sponsor. | Predicted positive amyloid PET status, assessed based on a in the Rotterdam Study validated prediction model (AUC = 0.84), including age and APOE ε4 allele count (0,1,2)^3^. |
| Body mass index (BMI) greater than 17 and less than 35 at Screening. | Body mass index > 17 and < 35, based on height and weight as measured at research visit. |
| If receiving an approved AD treatment, such as AChEIs, or memantine, or both for AD, must be on a stable dose for at least 12 weeks prior to Baseline. Treatment-naïve subjects for AD can be entered into the study. Unless otherwise stated, subjects must have been on stable doses of all other (ie, non-AD-related) permitted concomitant medications for at least 4 weeks prior to Baseline. | Not available in the Rotterdam Study. |
| Have an identified study partner (defined as a person able to support the subject for the duration of the study and who spends at least 8 hours per week with the subject). The study partner must provide separate written informed consent. In addition, this person must be willing and able to provide follow-up information on the subject throughout the course of the study. This person must, in the opinion of the investigator, spend sufficient time with the subject on a regular basis such that the study partner can reliably fulfill the study requirements. A permanent study partner need not be living in the same residence with the subject. For such a study partner not residing with the subject, the investigator has to be satisfied that the subject can contact the study partner readily during the times when the study partner is not with the subject. If in doubt about whether a subject's care arrangements are suitable for inclusion, the investigator should discuss this with the medical monitor. Study partners need to participate in person for visits where clinical assessment of CDR (global and CDR-SB), EQ-5D-5L, QOL-AD, ADCS MCI-ADL, and Zarit Burden Interview take place. | Has sufficient social support, as assessed based on a questionnaire “Do you have someone who can give you help when needed?”, answered with “yes”. Answers “no” and “maybe” were defined as insufficient social support. |
| Provide written informed consent. If a subject lacks capacity to consent in the investigator's opinion, the subject's assent should be obtained, if required in accordance with local laws, regulations and customs, plus the written informed consent of a legal representative should be obtained (capacity to consent and definition of legal representative should be determined in accordance with applicable local laws and regulations). In countries where local laws, regulations, and customs do not permit subjects who lack capacity to consent to participate in this study, they will not be enrolled. | All participants provided informed consent and authorization to partake in the Rotterdam Study and to monitor medical records, all participants were in full ability to understand the purpose and risks of the study. |
| Willing and able to comply with all aspects of the protocol. | All participants provided informed consent and authorization to partake in the Rotterdam Study and to monitor medical records, all participants were in full ability to understand the purpose and risks of the study. |

| **Clinical Trial Exclusion Criteria** | **Operational Study Definition** |
| --- | --- |
| Females who are breastfeeding or pregnant at Screening or Baseline (as documented by a positive beta-human chorionic gonadotropin [β-hCG] or human chorionic gonadotropin [hCG] test with a minimum sensitivity of 25 IU/L or equivalent units of β-hCG [or hCG]). A separate baseline assessment is required if a negative screening pregnancy test was obtained more than 72 hours before the first dose of study drug. | All women included were aged 50 years or older and therefore are presumed not to be at childbearing potential. Criteria for men were not available in the Rotterdam Study. |
| Females of childbearing potential who: Within 28 days before study entry, did not use a highly effective method of contraception, which includes any of the following:   - Total abstinence (if it is their preferred and usual lifestyle). - An intrauterine device or intrauterine hormone-releasing system (IUS). - A contraceptive implant. - An oral contraceptive (with additional barrier method) (Subject must be on a stable dose of the same oral contraceptive product for at least 28 days before dosing and throughout the study and for 28 days after study drug discontinuation.). - Have a vasectomized partner with confirmed azoospermia. - Do not agree to use a highly effective method of contraception (as described above) throughout the entire study period and for 28 days after study drug discontinuation. For sites outside of the EU, it is permissible that if a highly effective method of contraception is not appropriate or acceptable to the subject, then the subject must agree to use a medically acceptable method of contraception, ie, double-barrier methods of contraception such as latex or synthetic condomplus diaphragm or cervical/vault cap with spermicide. (amenorrheic for at least 12 consecutive months, in the appropriate age group, and without other known or suspected cause) or have been sterilized surgically (ie, bilateral tubal ligation, total hysterectomy, or bilateral oophorectomy, all with surgery at least 1 month before dosing). | All women included were aged 50 years or older and therefore are presumed not to be at childbearing potential. Criteria for men were not available in the Rotterdam Study. |
| Any neurological condition that may be contributing to cognitive impairment above and beyond that caused by the participant's Alzheimer's disease. | Prevalent Parkinson’s disease and stroke at research center visit, assessed by interview and verified using medical records.  History of TIA within 12 months prior to research center visit, as assessed by interview and by continuous linkage with medical records.  Prevalent brain cancer at research center visit, assessed by medical records and through linkage with a nationwide registry of histo- and cytopathology in the Netherlands, PALGA.  Prevalent multiple sclerosis, as assessed by medical records.  Presence of epilepsy at research center visit, assessed based on a questionnaire “Did you experience an episode of epilepsy since the last research visit?”. |
| History of transient ischemic attacks (TIA), stroke, or seizures within 12 months of Screening. | History of stroke or TIA within 12 months prior to research center visit, as assessed by interview and by continuous linkage with medical records. |
| Any psychiatric diagnosis or symptoms (example, hallucinations, major depression, or delusions) that could interfere with study procedures in the participant. | Presence of major depressive disorder of bipolar disorder within 1 year prior to research center visit, based on the Dutch version of the Schedules for Clinical Assessment in Neuropsychiatry (SCAN) questionnaire, and monitoring through medical records^9^. |
| Geriatric Depression Scale (GDS) score ≥ 8 at Screening. | A center for epidemiologic studies depression scale (CES-D) ≥16, which has similar predictive validity compared to the GDS^10^. |
| Contraindications to MRI scanning, including cardiac pacemaker/defibrillator, ferromagnetic metal implants (example in skull and cardiac devices other than those approved as safe for use in MRI scanners). | Presence of a contraindication for MRI. |
| Evidence of other clinically significant lesions on brain MRI at Screening that could indicate a dementia diagnosis other than Alzheimer's disease. | No clear causes of abnormality that would suggest another potential pathology for progressive dementia on visual ratings of MRI, including brain tumor or multiple sclerosis^5^. |
| Other significant pathological findings on brain MRI at Screening, including but not limited to:   - more than 4 microhemorrhages (defined as 10 mm or less at the greatest diameter); - a single macrohemorrhage greater than 10 mm at greatest diameter; - an area of superficial siderosis; - evidence of vasogenic edema; - evidence of cerebral contusion, encephalomalacia, aneurysms, vascular malformations, or infective lesions; - evidence of multiple lacunar infarcts or stroke involving a major vascular territory, severe small vessel, or white matter disease; - space occupying lesions; - or brain tumors (however, lesions diagnosed as meningiomas or arachnoid cysts and less than 1 cm at their greatest diameter need not be exclusionary). | Brain MRI shows no evidence of ≥ 2 lacunar infarcts, cerebellar infarctions, cortical infarctions, > 4 microbleeds, intracranial aneurism, meningioma > 1cm, glioma’s, arachnoid cysts, cavernous angioma, fistula or superficial siderosis based on visual ratings. A detailed protocol of the Rotterdam Scan Study is described elsewhere^5^.  Fazekas score of 3 based on volumetric segmentations (> 16.1ml of white matter hyperintensities)^5^. |
| Hypersensitivity to BAN2401 or any of the excipients, or to any monoclonal antibody treatment. | Not available in the Rotterdam Study. |
| Any immunological disease which is not adequately controlled, or which requires treatment with biologic drugs during the study. | Use of immunosuppressant medication (ATC-code: L04A), based on pharmacy dispensing records. |
| Subjects with a bleeding disorder that is not under adequate control (including a platelet count < 50,000 or international normalized ratio [INR] > 1.5). | A platelet count of < 50,000, based on fasting blood samples. |
| Have thyroid stimulating hormone above normal range. Other tests of thyroid function with results outside the normal range should only be exclusionary if they are considered clinically significant by the investigator. This applies to all subjects whether or not they are taking thyroid supplements. | Not available in the Rotterdam Study. |
| Abnormally low serum vitamin B12 levels for the testing laboratory (if subject is taking vitamin B12 injections, level should be at or above the lower limit of normal [LLN] for the testing laboratory). Levels of Vitamin B12 may be confirmed with reflex testing to include methylmalonic acid (MMA) analysis, if available in region. | Not available in the Rotterdam Study. |
| Known to be human immunodeficiency virus (HIV) positive. | Not available in the Rotterdam Study. |
| Any other clinically significant abnormalities in physical examination, vital signs, laboratory tests, or ECG at Screening or Baseline which in the opinion of the principal investigator (PI) require further investigation or treatment or which may interfere with study procedures or safety. | Presence of second or third degree AV-block based on MEANS interpretation, assessed by research physicians using ECG from the research center visit^7^. |
| Subjects with malignant neoplasms within 3 years of Screening (except for basal or squamous cell carcinoma in situ of the skin, or localized prostate cancer in male subjects). Subjects who had malignant neoplasms but who have had at least 3 years of documented uninterrupted remission before Screening need not be excluded. | History of cancer (solid and hematological, with exclusion of non-melanoma skin cancer) within 3 years prior to research center visit, as assessed by medical records and through linkage with a nationwide registry of histo- and cytopathology in the Netherlands, PALGA. |
| Answer “yes” to Columbia-Suicide Severity Rating Scale (C-SSRS) suicidal ideation Type 4 or 5, or any suicidal behavior assessment within 6 months before Screening, at Screening, or at the Baseline Visit, or has been hospitalized or treated for suicidal behavior in the past 5 years before Screening. | Not available in the Rotterdam Study for participants with CES-D score < 16. |
| Known or suspected history of drug or alcohol abuse or dependence within 2 years before Screening or a positive urine drug test at Screening. Subjects who test positive for benzodiazepines or opioids in urine drug testing need not be excluded if in the clinical opinion of the investigator, this is due to the subject taking prior/concomitant medications containing benzodiazepines or opioids for a medical condition and not due to drug abuse. | For the fourth examination round research visits, the presence of alcohol abuse based on the CAGE questionnaire^6^.  For the fifth visits, assessed by a questionnaire “How many units of alcohol do you consume during one week?” answered with ≥ 6. |
| Any other medical conditions (example, cardiac, respiratory, gastrointestinal, renal disease) which are not stably and adequately controlled, or which in the opinion of the investigator(s) could affect the participant's safety or interfere with the study assessments. | Presence of renal disease at research center visit, as assessed by questionnaire “Are you receiving dialysis due to impaired kidney function?” or estimated glomerular filtration rate < 30 based on fasting blood samples.  During the fifth research center visit, positive test result screening for the presence of impaired liver function, assessed by aspartate aminotransferase and alanine aminotransferase levels > 90. Not available for the fourth research center visit.  History of heart failure, coronary heart disease (including myocardial infarction, percutaneous coronary angioplasty and coronary artery bypass grafting) or incident atrial fibrillation within 12 months prior to research center visit, monitored using medical records. |
| Subjects who are taking prohibited medications. | Not available in the Rotterdam Study. |
| Participation in a clinical study involving any therapeutic monoclonal antibody, protein derived from a monoclonal antibody, immunoglobulin therapy, or vaccine within 6 months before screening unless it can be documented that the participant was randomized to placebo. | Not available in the Rotterdam Study. |
| Subjects who have any known prior exposure to BAN2401. | Not available in the Rotterdam Study. |
| Subjects who were dosed in a clinical study involving any new chemical entities for AD within 6 months prior to Screening unless it can be documented that the subject was in a placebo treatment arm. | Not available in the Rotterdam Study. |
| Participated in any other investigational medication or device study in the 8 weeks or 5 half-lives (whichever is longer) of the medication before randomization unless it can be documented that the subject was in a placebo treatment arm. | Not available in the Rotterdam Study. |
| Planned surgery which requires general anesthesia that would take place during the study. Planned surgery which requires only local anesthesia and which can be undertaken as day case without inpatient stay postoperatively need not result in exclusion if in the opinion of the PI this operation does not interfere with study procedures and subject safety. | Not available in the Rotterdam Study. |
| Severe visual or hearing impairment that would prevent the subject from performing psychometric tests accurately. | By design, all participants included have adequate literacy, vision and hearing for neuropsychological testing. |

| **Supplementary table 4. TRAILBLAZER-ALZ2 (donanemab) Clinical Trial Inclusion and Exclusion Criteria and Operationalization in the Rotterdam Study** | |
| --- | --- |
| **Clinical Trial Inclusion Criteria** | **Operational Study Definition** |
| 60 to 85 years of age inclusive, at the time of signing the informed consent. | Age between 60 and 85 years at research center visit, based on date of birth municipality data. |
| MMSE score of 20 to 28 (inclusive) at baseline (Visit 601 or 1.). | MMSE score of ≥ 20 and ≤ 28 assessed during home interview. |
| Gradual and progressive change in memory function reported by the participant or informant for ≥ 6 months. | Diagnosis of MCI at study center visit, as defined as presence of i) subjective cognitive complaints, ii) objective cognitive impairment, and iii) absence of dementia. Details of MCI diagnosis in the Rotterdam Scan Study is described^11^.    Diagnosis of AD within a year prior or after research center visit according to NINCDS-ADRDA criteria, continuously monitored through linkage with computerized medical records and discussed through a panel with an expert neurologist^2^. |
| Meet 18F flortaucipir PET scan (central read) criteria. | Predicted positive amyloid PET status, assessed based on a in the Rotterdam Study validated prediction model (AUC = 0.84), including age and APOE ε4 allele count (0,1,2)^3^. |
| Have a study partner who will provide written informed consent to participate, is in frequent contact with the participant (defined as at least 10 hours per week), and will accompany the participant to study visits or be available by telephone at designated times. A second study partner may serve as backup. The study partner(s) is/are required to accompany the participant for signing consent. One study partner is requested to be present or available by phone on all days the C-SSRS/Self-Harm Supplement Form is administered. The study partner must be present on all days the cognitive and functional scales are administered. If a participant has a second study partner, it is preferred that 1 study partner be primarily responsible for the CDR and Alzheimer’s Disease Cooperative Study - Activities of Daily Living Inventory (ADCS-ADL) assessments. Visits requiring the following assessments and scales must have a study partner available by telephone if not accompanying participant at a visit for the following assessments: AEs and concomitant medications Relevant portions of the C-SSRS/Self-Harm Supplement Forms CDR, and ADCS-ADL. If a study partner must withdraw from study participation, a replacement may be allowed at the investigator’s discretion. The replacement will need to sign a separate informed consent on the first visit that he or she accompanies the participant. | Has sufficient social support, as assessed based on a questionnaire “Do you have someone who can give you help when needed?”, answered with “yes”. Answers “no” and “maybe” were defined as insufficient social support. |
| Have adequate literacy, vision, and hearing for neuropsychological testing in the opinion of the investigator at the time of screening. | By design, all participants included have adequate literacy, vision and hearing for neuropsychological testing. |
| Are reliable and willing to make themselves available for the duration of the study and are willing to follow study procedures. | All participants provided informed consent and authorization to partake in the Rotterdam Study and to monitor medical records, all participants were in full ability to understand the purpose and risks of the study. |
| Stable concomitant symptomatic AD medications and other medication that may impact cognition for at least approximately 30 days prior to randomization (does not apply to topical, as needed [prn], or discontinued medications). | Not available in the Rotterdam Study. |
| Males and females will be eligible for this study.  Contraceptive use by men or women should be consistent with local regulations regarding the methods of contraception for those participating in clinical studies. b. Male participants: i. Men, regardless of their fertility status, with non-pregnant women of childbearing potential (WOCBP) partners must agree to either remain abstinent (if this is their preferred and usual lifestyle) or use condoms as well as 1 additional highly effective (less than 1% failure rate) method of contraception (such as combination oral contraceptives, implanted contraceptives, or intrauterine devices) or effective method of contraception (such as diaphragms with spermicide or cervical sponges) for the duration of the study and until their plasma concentrations are below the level that could result in a relevant potential exposure to a possible fetus, predicted to be 90 days following last dose of IP. A. Men and their partners may choose to use a double-barrier method of contraception. (Barrier protection methods without concomitant use of a spermicide are not an effective or acceptable method of contraception. Thus, each barrier method must include use of a spermicide. It should be noted, however, that the use of male and female condoms as a double barrier method is not considered acceptable due to the high failure rate when these barrier methods are combined). B. Periodic abstinence (e.g., calendar, ovulation, symptothermal, post-ovulation methods), declaration of abstinence just for the duration of a trial, and withdrawal are not acceptable methods of contraception. ii. Men with pregnant partners should use condoms during intercourse for the duration of the study and until the end of estimated relevant potential exposure in WOCBP (90 days). iii. Men should refrain from sperm donation for the duration of the study and until their plasma concentrations are below the level that could result in a relevant potential exposure to a possible fetus, predicted to be 90 days following last dose of IP. iv. Men who are in exclusively same sex relationships (as their preferred and usual lifestyle) are not required to use contraception. c. Female participants: i. Women not of childbearing potential may participate and include those who are: A. infertile due to surgical sterilization (hysterectomy, bilateral oophorectomy, or tubal ligation), congenital anomaly such as Mullerian agenesis; or B. post-menopausal - defined as either a. A woman at least 40 years of age with an intact uterus, not on hormone therapy, who has cessation of menses for at least 1 year without an alternative medical cause, AND a follicle-stimulating hormone >40 mIU/mL; or b. A woman 55 or older not on hormone therapy, who has had at least 12 months of spontaneous amenorrhea; or c. A woman at least 55 years of age with a diagnosis of menopause prior to starting hormone replacement therapy. | All women included were aged 60 years or older and are presumed not to be at childbearing potential. Criteria for men were not available in the Rotterdam Study. |
| Capable of giving signed informed consent as described in Section 10.1.3 which includes compliance with the requirements and restrictions listed in the informed consent form (ICF) and in this protocol. | All participants provided informed consent and authorization to partake in the Rotterdam Study and to monitor medical records, all participants were in full ability to understand the purpose and risks of the study. |

| **Clinical Trial Exclusion Criteria** | **Operational Study Definition** |
| --- | --- |
| Significant neurological disease affecting the central nervous system other than AD, that may affect cognition or ability to complete the study, including but not limited to, other dementias, serious infection of the brain, Parkinson’s disease, multiple concussions or epilepsy or recurrent seizures (except febrile childhood seizures). | Prevalent Parkinson’s disease and stroke at research center visit, as assessed by interview and verified using medical records.  History of TIA within 12 months prior to research center visit, as assessed by interview and by continuous linkage with medical records.  Prevalent brain cancer at research center visit, as assessed by medical records and through linkage with a nationwide registry of histo- and cytopathology in the Netherlands, PALGA.  Prevalent multiple sclerosis, as assessed by medical records.  Presence of epilepsy at research center visit, as assessed based on a questionnaire “Did you experience an episode of epilepsy since the last research visit?”. |
| Current serious or unstable illnesses including cardiovascular, hepatic, renal, gastroenterologic, respiratory, endocrinologic, neurologic (other than AD), psychiatric, immunologic, or hematologic disease and other conditions that, in the investigator’s opinion could interfere with the analyses in this study; or has a life expectancy of < 24 months. | History of heart failure or coronary heart disease (including myocardial infarction, percutaneous coronary angioplasty, coronary artery bypass grafting) within 2 years prior to research center visit, monitored using medical records.  Presence of renal disease at research center visit, as assessed by questionnaire “Are you receiving dialysis due to impaired kidney function?”.  During the fifth research center visit, positive test result screening for hepatitis C virus antibody or hepatitis B virus, based on level of ≥ 1.0 in the Elecsys Anti-HCV II and the HbsAg immunoassays^8^. Not available for the fourth research center visit.  Presence of abnormal thyroid function at home visit, as assessed based on a questionnaire “Since the last research visit, did you experience any disease concerning the thyroid that needed treatment from the general practitioner or a specialist?”.  Presence of major depressive disorder of bipolar disorder within 6 months prior to research center visit, based on the Dutch version of the SCAN questionnaire, and monitoring through medical records^4^.  History of deep vein thrombosis or pulmonary embolism, as assessed by questionnaires during home interview: “Since the last research visit, did you experience deep vein thrombosis of the leg?” and “Since the last research visit, did you experience pulmonary embolism?”. |
| History of cancer within the last 5 years, with the exception of non-metastatic basal and/or squamous cell carcinoma of the skin, in situ cervical cancer, nonprogressive prostate cancer, or other cancers with low risk of recurrence or spread. | History of cancer (solid and hematological, with exclusion of non-melanoma skin cancer) within 5 years prior to research center visit, as assessed by medical records and through linkage with a nationwide registry of histo- and cytopathology in the Netherlands, PALGA. |
| Participants with any current primary psychiatric diagnosis other than AD if, in the judgment of the investigator, the psychiatric disorder or symptom is likely to confound interpretation of drug effect, affect cognitive assessment, or affect the participant’s ability to complete the study. Participants with history of schizophrenia or other chronic psychosis are excluded. | Presence of major depressive disorder of bipolar disorder at research center visit, based on the Dutch version of the SCAN questionnaire.  Presence of Anxiety at research center visit, based on the Composite International Diagnostic Interview^12^. |
| Are, in the judgment of the investigator, actively suicidal and therefore deemed to be at significant risk for suicide. | Not available in the Rotterdam Study. |
| History of alcohol or drug use disorder (except tobacco use disorder) within 2 years before the screening visit. | For the fourth examination round research visits, the presence of alcohol abuse based on the CAGE questionnaire^6^.  For the fifth visits, assessed by a questionnaire “How many units of alcohol do you consume during one week?” answered with ≥ 6. |
| History of clinically significant multiple or severe drug allergies, significant atopy, or severe posttreatment hypersensitivity reactions (including but not limited to erythema multiforme major, linear immunoglobulin A dermatosis, toxic epidermal necrolysis, and/or exfoliative dermatitis). | Not available in the Rotterdam Study. |
| Have any clinically important abnormality at screening, as determined by investigator, in physical or neurological examination, vital signs, ECG, or clinical laboratory test results that could be detrimental to the participant, could compromise the study, or show evidence of other etiologies for dementia. | Presence of atrial fibrillation, myocardial infarction or second or third degree AV-block based on MEANS interpretation, as assessed by ECG performed at the research center visit^7^. |
| Screening MRI which shows evidence of significant abnormality that would suggest another potential etiology for progressive dementia or a clinically significant finding that may impact the participant’s ability to safely participate in the study. | No clear causes of abnormality that would suggest another potential pathology for progressive dementia on visual ratings of MRI, including brain tumor or multiple sclerosis^5^. |
| Have any contraindications for MRI, including claustrophobia or the presence of contraindicated metal (ferromagnetic) implants/cardiac pacemaker. | Presence of contraindication for MRI. |
| Have a centrally read MRI demonstrating presence of ARIA-E, > 4 cerebral microhemorrhages, more than 1 area of superficial siderosis, any macrohemorrhage or severe white matter disease at screening. | Brain MRI shows no evidence of > 4 microhaemorrhages (< 10mm) or superficial siderosis based on visual ratings. A detailed protocol of the Rotterdam Scan Study is described elsewhere^5^.  Fazekas score of 3 based on volumetric segmentations (> 16.1ml of white matter hyperintensities)^5^. |
| Sensitivity to florbetapir F18 or flortaucipir F18. | Not available in the Rotterdam Study. |
| Poor venous access. | Not available in the Rotterdam Study. |
| Contraindication to PET. | Not available in the Rotterdam Study. |
| Present or planned exposure to ionizing radiation that, in combination with the planned administration of study PET ligands, would result in a cumulative exposure that exceeds local recommended exposure limits. | Not available in the Rotterdam Study. |
| Alanine aminotransaminase (ALT) ≥ 2.5X the upper limit of normal (ULN) of the performing laboratory, aspartate aminotransferase (AST) ≥ 2.5X ULN, total bilirubin level (TBL) ≥ 1.5X ULN, or alkaline phosphatase (ALP) ≥ 2X ULN at screening.  Note: participant with TBL ≥ 1.5X ULN are not excluded if they meet all of the following criteria for Gilbert syndrome: Bilirubin is predominately indirect (unconjugated) at screening (direct bilirubin within normal limits). Absence of liver disease. ALT, AST, and ALP ≤1X ULN at screening.  Hemoglobin is not significantly decrease at screening. | Fasting blood sample alanine aminotransaminase or aspartate aminotransferase levels > 112,5, based on an upper limit of normal of 45.  Fasting blood sample total bilirubin levels of > 25.5, based on an upper limit of normal of 17.  Fasting blood sample alkaline phosphatase levels > 268, based on an upper limit of normal of 134.  Not available for the fourth research center visit. |
| Have had prior treatment with a passive anti-amyloid immunotherapy. | Not available in the Rotterdam Study. |
| Have received active immunization against Abeta in any other study. | Not available in the Rotterdam Study. |
| Have known allergies to donanemab, related compounds, or any components of the formulation. | Not available in the Rotterdam Study. |
| Are currently enrolled in any other interventional clinical trial involving an IP or any other type of medical research judged not to be scientifically or medically compatible with this study. | Not available in the Rotterdam Study. |
| Have participated, within the last 30 days (4 months for studies conducted in Japan; 3 months for studies conducted in the United Kingdom), in a clinical trial involving an IP. If the previous IP is scientifically or medically incompatible with this study and has a long half-life, 3 months or 5 half-lives (whichever is longer) should have passed prior to screening (participation in observational studies may be permitted upon review of the observational study protocol and approval by the sponsor). | Not available in the Rotterdam Study. |
| Have previously completed or withdrawn from this study or received donanemab in any prior investigational study. (This exclusion criterion does not apply to participants who are allowed to rescreen before randomization in this study). | Not available in the Rotterdam Study. |
| Are investigator site personnel directly affiliated with this study and/or their immediate families. Immediate family is defined as a spouse, parent, child, or sibling, whether biological or legally adopted. | Not available in the Rotterdam Study. |
| Are Lilly employees or are employees of third-party organizations (TPOs) involved in study which requires exclusion of their employees, or have study partners who are Lilly employees or are employees of TPOs involved in a study which requires exclusion of their employees. | Not available in the Rotterdam Study. |

**Supplementary figure 1. Flowchart of participants selected for analysis**

**All participants**

4^th^ examination round, N = 6,052

5^th^ examination round, N = 7,162

Participants with MCI, N = 779

Participants with early AD, N = 189

**Aducanumab**

MMSE score ≥ 24

MCI, N = 667

Early AD, N = 84

**Lecanemab**

MMSE score ≥ 22

MCI, N = 748

Early AD, N = 84

**Donanemab**

MMSE score 22-28

MCI, N = 650

Early AD, N = 112

**Supplementary table 5. Prevalence of Trial Eligibility Criteria among Participants with Mild Cognitive Impairment in the Population-based Rotterdam Study**

|  | Aducanumab^1^ | Lecanemab^2^ | Donanemab^3^ |
| --- | --- | --- | --- |
|  | N = 751 | N = 856 | N = 762 |
| Age range | 67 (8.9) | 17 (2.0) | 94 (12.3) |
| Predicted amyloid negative | 318 (42.3) | 346 (40.4) | 289 (37.9) |
| No social support | 113 (15.0) | 134 (15.6) | 120 (15.7) |
| Impaired 15-word learning test | n/a | 391 (46.7) | n/a |
| Body mass index | n/a | 43 (4.9) | n/a |
| Anticoagulant medication use | 234 (31.2) | n/a | n/a |
| Other medication use | 153 (20.4) | 7 (0.8) | n/a |
| Cardiovascular disease | 264 (35.2) | 73 (8.5) | 73 (9.6) |
| Neurological disease | 103 (13.7) | 116 (13.2) | 112 (14.7) |
| Psychiatric disorder | 20 (2.7) | 138 (16.1) | 121 (15.9) |
| Clotting disorder | 10 (1.3) | - | n/a |
| Cancer | 46 (6.1) | 52 (5.9) | 57 (7.5) |
| Thyroid dysfunction | 40 (5.3) | n/a | 38 (5.0) |
| Hepatological disorder | 7 (0.9) | 2 (0.2) | 18 (2.4) |
| Impaired renal function | n/a | 7 (0.8) | n/a |
| Any clinical criterion | 573 (76.3) | 649 (75.8) | 456 (59.8) |
| Any clinical criterion + predicted amyloid negativity | 664 (88.4) | 736 (86.0) | 582 (76.4) |

All variables are expressed as N (%). n/a indicates not applicable. ^1^ Eligibility criteria as applied in EMERGE trial. ^2^ Eligibility criteria as applied in CLARITY-AD trial. ^3^ Eligibility as applied in TRALBLAZER-ALZ2 trial. Other medication for aducanumab includes urological medication, corticosteroids, immunosuppressant medication, anti-epileptic medication, benzodiazepines, antipsychotic medication and tricyclic antidepressants. Other medication for lecanemab includes immunosuppressant medication.

**Supplementary Table 6. Prevalence of Trial Eligibility Criteria among Participants with Mild Cognitive Impairment, age within the trial age range and predicted positive amyloid status in the Population-based Rotterdam Study**

|  | Aducanumab^1^ | Lecanemab^2^ | Donanemab^3^ |
| --- | --- | --- | --- |
|  | N = 282 | N = 391 | N = 295 |
| No social support | 52 (18.4) | 72 (18.4) | 45 (15.3) |
| Impaired 15-word learning test | n/a | 182 (46.5) | n/a |
| Body mass index | n/a | 17 (4.3) | n/a |
| Anticoagulant medication use | 86 (30.5) | n/a | n/a |
| Other medication use | 49 (17.3) | 2 (0.5) | n/a |
| Cardiovascular disease | 97 (34.4) | 42 (10.7) | 25 (8.5) |
| Neurological disease | 39 (13.8) | 48 (12.3) | 42 (14.2) |
| Psychiatric disorder | 8 (2.8) | 62 (17.5) | 42 (14.2) |
| Clotting disorder | 6 (2.1) | n/a | n/a |
| Cancer | 15 (5.3) | 20 (5.1) | 19 (6.4) |
| Thyroid dysfunction | 16 (5.7) | n/a | 16 (5.4) |
| Hepatological disorder | 1 (0.4) | 0 (0.0) | 3 (1.0) |
| Impaired renal function | n/a | 4 (1.0) | n/a |
| Any clinical criterion | 208 (73.8) | 282 (72.1) | 146 (49.5) |

All variables are expressed as N (%). n/a indicates not applicable. ^1^ Eligibility criteria as applied in EMERGE trial. ^2^ Eligibility criteria as applied in CLARITY-AD trial. ^3^ Eligibility as applied in TRALBLAZER-ALZ2 trial. Other medication for aducanumab includes urological medication, corticosteroids, immunosuppressant medication, anti-epileptic medication, benzodiazepines, antipsychotic medication and tricyclic antidepressants. Other medication for lecanemab includes immunosuppressant medication.

**Supplementary table 7. Prevalence of Aducanumab exclusion criteria per individual criterion**

|  | | Aducanumab^1^ | | | Lecanemab^2^ | | Donanemab^3^ | | |
| --- | --- | --- | --- | --- | --- | --- | --- | --- | --- |
|  | | 4^th^ examination round | 5^th^ examination round | 4^th^ examination round | | 5^th^ examination round | | 4^th^ examination round | 5^th^ examination round |
|  |  | N = 430 | N = 321 | N = 509 | | N = 347 | | N = 461 | N = 301 |
| Visit age, mean (SD) | | 73.52 (7.71) | 74.54 (8.24) | 74.16 (7.91) | | 74.80 (8.35) | | 75.23 (8.00) | 75.28 (8.26) |
| Female sex | | 223 (51.9) | 165 (51.4) | 281 (55.2) | | 180 (51.9) | | 261 (56.6) | 162 (53.8) |
| Mini-mental state exam score, mean (SD) | | 26.83 (1.68) | 27.10 (1.57) | 26.16 (2.20) | | 26.73 (1.96) | | 25.16 (2.25) | 25.84 (2.03) |
| Age outside trial range | | 33 (7.7) | 34 (10.6) | 10 (2.0) | | 7 (2.0) | | 58 (12.6) | 36 (12.0) |
| Predicted amyloid positive | | 235 (54.7) | 198 (61.7) | 288 (56.6) | | 222 (64.0) | | 279 (60.5) | 194 (64.5) |
| No social support | | 80 (18.6) | 33 (10.3) | 94 (18.5) | | 46 (13.3) | | n/a | n/a |
| Unimpaired 15-word learning test | | n/a | n/a | 239 (47.0) | | 152 (43.8) | | n/a | n/a |
| Body mass index <17 or >35 | | n/a | n/a | 20 (3.9) | | 23 (6.6) | | 83 (18.0) | 37 (12.3) |
| Multiple sclerosis | | 2 (0.5) | 0 (0.0) | 3 (0.6) | | 1 (0.3) | | 3 (0.7) | 1 (0.3) |
| Transient ischemic attack | | 0 (0.0) | 4 (1.2) | 0 (0.0) | | 4 (1.2) | | 3 (0.7) | 4 (1.3) |
| Stroke | | 42 (9.8) | 30 (9.3) | 49 (9.6) | | 32 (9.2) | | 48 (10.4) | 30 (10.0) |
| Epilepsy | | 18 (4.2) | 6 (4.8) | 20 (3.9) | | 6 (4.3) | | 17 (3.7) | 8 (6.2) |
| Parkinson | | 5 (1.2) | 1 (0.3) | 5 (1.0) | | 1 (0.3) | | 4 (0.9) | 0 (0.0) |
| Heart failure | | 7 (1.6) | 2 (0.6) | 7 (1.4) | | 2 (0.6) | | 8 (1.7) | 3 (1.0) |
| Coronary heart disease | | 5 (1.2) | 3 (0.9) | 5 (1.0) | | 3 (0.9) | | 6 (1.3) | 4 (1.3) |
| Atrial fibrillation | | 3 (0.7) | 4 (1.2) | 3 (0.6) | | 4 (1.2) | | 3 (0.7) | 4 (1.3) |
| Hypertension | | 105 (24.4) | 67 (20.9) | n/a | | n/a | | n/a | n/a |
| Uncontrolled diabetes mellitus | | 37 (8.6) | 30 (9.3) | n/a | | n/a | | n/a | n/a |
| Bundle branch block electrocardiogram | | 24 (5.6) | 23 (7.2) | 28 (5.8) | | 25 (7.2) | | 28 (6.1) | 23 (7.6) |
| Low platelet count | | n/a | n/a | 1 (0.2) | | 0 (0.0) | | n/a | n/a |
| Depression | | 11 (2.6) | n/a | 14 (2.8) | | n/a | | 14 (3.0) | NA |
| Bipolar disorder | | 2 (0.5) | n/a | 2 (0.4) | | n/a | | 2 (0.4) | NA |
| Center of epidemiologic studies depression scale ≥ 16 | | n/a | n/a | 80 (15.7) | | 49 (14.1) | | n/a | n/a |
| Alcohol abuse | | 4 (0.9) | 3 (0.9) | 5 (1.0) | | 3 (0.9) | | 4 (0.9) | 2 (0.7) |
| Anxiety disorder | | n/a | n/a | n/a | | n/a | | 55 (11.9) | 19 (6.3) |
| Clinical Assessment in Neuropsychiatry depression | | n/a | n/a | n/a | | n/a | | 45 (9.8) | 18 (6.0) |
| Anticoagulants | | 130 (30.2) | 104 (32.4) | n/a | | n/a | | n/a | n/a |
| Urological medication | | 0 (0.0) | 2 (0.6) | n/a | | n/a | | n/a | n/a |
| Corticosteroids | | 6 (1.4) | 8 (2.5) | n/a | | n/a | | n/a | n/a |
| Immunosuppressant medication | | 0 (0.0) | 7 (2.2) | 0 (0.0) | | 7 (2.0) | | n/a | n/a |
| Opioid mediation | | 6 (1.4) | 12 (3.7) | n/a | | n/a | | n/a | n/a |
| Anti-epileptic medication | | 15 (3.5) | 9 (2.8) | n/a | | n/a | | n/a | n/a |
| Benzodiazepines | | 63 (14.7) | 30 (9.3) | n/a | | n/a | | n/a | n/a |
| Antipsychotic medication | | 6 (1.4) | 3 (0.9) | n/a | | n/a | | n/a | n/a |
| Tricyclic antidepressants | | 15 (3.5) | 8 (2.5) | n/a | | n/a | | n/a | n/a |
| History of thrombosis | | 3 (0.7) | 2 (0.6) | n/a | | n/a | | n/a | n/a |
| History of lung embolus | | 5 (1.2) | 1 (0.3) | n/a | | n/a | | n/a | n/a |
| Hepatological disorder | | n/a | 7 (2.2) | n/a | | 2 (0.6) | | n/a | 18 (6.0) |
| Cancer | | 22 (5.1) | 24 (7.5) | 27 (5.3) | | 25 (7.2) | | 32 (6.9) | 25 (8.3) |
| Thyroid dysfunction | | 29 (6.7) | 11 (3.4) | n/a | | n/a | | 27 (5.9) | 11 (3.7) |
| Impaired renal function | | n/a | n/a | 3 (0.6) | | 3 (0.9) | | n/a | n/a |
| Dialysis | | 0 (0.0) | 0 (0.0) | 47 (9.2) | | 0 (0.0) | | 0 (0.0) | 0 (0.0) |

All covariates are expressed as N (%) unless specified otherwise. n/a indicates not applicable.

Aducanumab^1^: Table contains imputed data, we had no missing values for age, multiple sclerosis, epilepsy, stroke, heart failure, atrial fibrillation, anticoagulant and other medication, cancer and dialysis. N (%) missing for missing variables predicted amyloid status 20 (2.7%), social support 130 (17.9%), Parkinson 47 (7.0%), transient ischemic attack 43 (6.4%), coronary heart disease 6 (0.9%), hypertension 44 (6.6%), uncontrolled diabetes mellitus 99 (14.8%), bundle branch block 56 (8.3%), depression 4^th^ examination round 6 (0.9%), bipolar disorder 6 (0.9%), alcohol abuse 3 (0.5%), thyroid dysfunction 4 (0.6%), thrombosis leg 216 (32.2%), lung embolus 216 (32.2%), hepatological dysfunction 5^th^ examination round 46 (16.0%).

Lecanemab^2^: Table contains imputed data, we had no missing values for age, multiple sclerosis, stroke, heart failure, atrial fibrillation, immunosuppressant medication, cancer and dialysis. N (%) missing for missing variables predicted amyloid status 36 (4.2%), social support 132 (15.4%), body mass index 63 (7.4%), 15-word learning test 28 (3.3%), epilepsy 1 (0.01%), Parkinson 54 (6.3%), transient ischemic attack 47 (5.5%), coronary heart disease 6 (0.7%), bundle branch block 64 (7.5%), depression 4^th^ examination round 8 (1.6%), bipolar disorder 8 (1.6%), alcohol abuse 4 (0.5%), impaired renal function 97 (11.3%), hepatological dysfunction 5^th^ examination round 62 (19.3%).

Donanemab^3^: Table contains imputed data, we had no missing values for age, multiple sclerosis, stroke, heart failure, atrial fibrillation, cancer and dialysis. N (%) missing for missing variables predicted amyloid status 21 (2.8%), social support 115 (15.1%), epilepsy 1 (0.01%), Parkinson 45 (5.9%), transient ischemic attack 40 (5.2%), coronary heart disease 5 (0.7%), bundle branch block 55 (7.2%), depression 4^th^ examination round 10 (1.3%), bipolar disorder 10 (1.3%), clinical assessment in neuropsychiatry depression 3 (0.4%), anxiety disorder 39 (9.1%), alcohol abuse 4 (0.5%), thyroid dysfunction 1 (0.01%), hepatological dysfunction 5^th^ examination round 46 (15.2%).

3.	Nguyen Ho PT, van Arendonk, J., Steketee, R. M., van Rooij, F. J., Roshchupkin, G. V., Ikram, M. A., ... & Neitzel, J. . Predicting amyloid‐beta pathology in the general population. *Alzheimer's & Dementia*. 2023;

4.	United Nations Educational SaCOU. International Standard Classification of Education (ISCED). 1976;
